# Development and Evaluation of an Offline Vision–Language Model System for Estimating Nutrients from Meal Photographs on Personal Devices: The PLANS (Personal-device Local Assessment of Nutrients System) Study

**DOI:** 10.64898/2026.10.11.26365273

**Authors:** Shinichi Nakagawa, Akira Yamamoto

## Abstract

**Background:** Estimating nutrient intake from meal photographs with vision–language models (VLMs) is now practical, but patient photographs often cannot leave the institution. We developed PLANS (Personal-device Local Assessment of Nutrients System), in which a small open VLM runs entirely on a personal device.

**Methods:** Open VLMs were compared on two image sets with reference values (NutriImage, 887 Japanese meals; SNAPMe, 1,134 US meals), and the selected model (Qwen3-VL-4B, 4-bit) was run on an iPad Air without network access. Energy (primary outcome), protein, fat, carbohydrate and salt were evaluated by median absolute percentage error (MedAPE) and Spearman’s ρ. The effect of supplying the food weight, including errors in it, was examined.

**Results:** Energy estimates on the iPad were identical to those of a desktop-computer run for 84–86% of images (other values, 75–87%). MedAPE on the iPad was 37.0% and 36.5% for energy (26–28% of images within ±20%), 43–65% for the macronutrients and 68–101% for salt (ρ 0.51–0.85). On the desktop computer, supplying the true weight reduced MedAPE of the 4B model on NutriImage from 37.0% to 32.5% for energy and from 100.0% to 66.8% for salt; energy MedAPE stayed below the base value for declared weights of 0.5–1.25 times the true weight or rounded to 50 g, but rose markedly when the container weight was included.

**Conclusions:** An offline VLM on a tablet reproduced server-side energy accuracy and is a candidate tool for dietary counselling where photographs cannot leave the institution; given the wide limits of agreement, its usefulness in practice remains to be examined.

## 1. Introduction

Photograph-based dietary assessment could support the nutritional management of diabetes and dyslipidaemia without the burden of weighed records. Commercial vision–language models (VLMs) estimate energy from a single meal photograph with median absolute percentage errors of roughly 30–40% [1], but many medical institutions cannot send patients’ photographs to an external service for reasons of privacy and governance. The constraint is the transmission, not the model, and it applies to any image. A system in which a VLM runs entirely on a device within the institution, or on the patient’s own device, would remove this barrier, and meal photographs are one concrete task on which such a system can be tested.

Small open VLMs of 2–8 billion parameters can now run on personal computers and tablets, and offline calorie-tracking applications using a 2B-class VLM have appeared [2]. How the design choices of such a system—model size, numerical precision, the inference library and the information requested from the user—affect accuracy, and whether server-side accuracy carries over to the device, has not been examined systematically.

In previous work on Japanese and US meal images with reference values we observed that portion-related content in the prompt shifts the estimate systematically [1]. This suggests that the portion is inferred partly from what the dish is taken to be; if so, supplying information that the image cannot provide—the weight of the food—may help.

We describe the development and evaluation of PLANS (Personal-device Local Assessment of Nutrients System), an offline system for estimating energy and nutrients from meal photographs on personal Apple devices. We (i) selected a model from server-side comparisons of open VLMs; (ii) examined whether the selected model, quantized to 4 bits, reproduced server-side accuracy on a desktop computer and a tablet; (iii) examined the effect of supplying the food weight as text and its robustness to errors in the declared weight; and (iv) placed the result alongside current commercial models as a reference. Energy was the primary outcome; protein, fat, carbohydrate and salt equivalent were secondary. This is a feasibility and design study; photographs taken by users were not evaluated.

## 2. Materials and Methods

### 2.1. Study design

The study had four stages (Online Resource 1, Fig. S1): (1) comparison of open VLM families and sizes on a server; (2) reproduction of the selected model at 4 bits on a desktop computer and a tablet; (3) evaluation of the food weight supplied as text, including errors in the declared weight; and (4) runs of current commercial models as an accuracy reference. All inferences used the same base prompt and image files, with temperature 0 for open models. The persona conditions of ref. [1] were run only to check whether a model’s response to prompt content survived quantization; they are not part of PLANS (Fig. 1).

**Fig 1.**
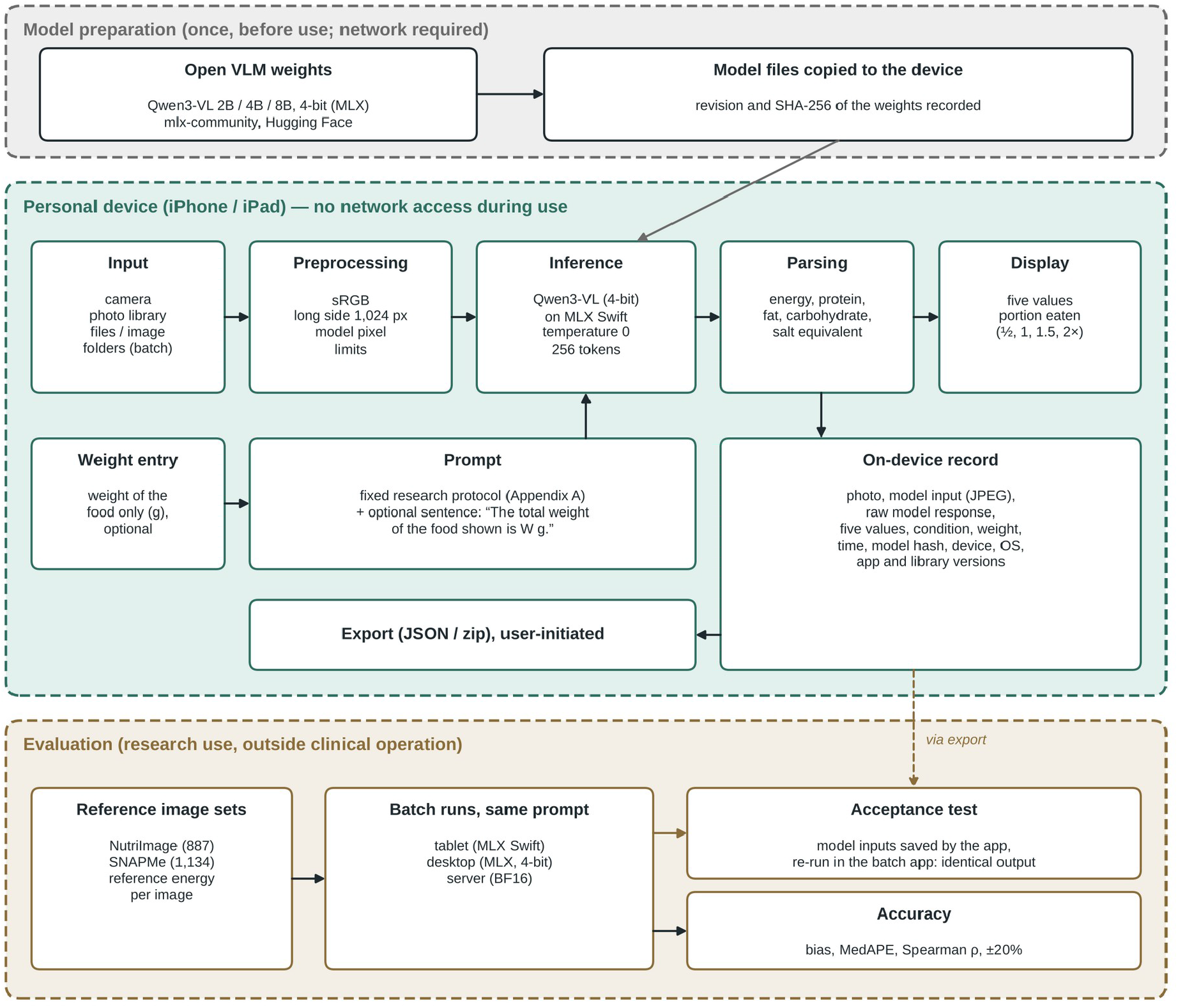
Overview of PLANS (Personal-device Local Assessment of Nutrients System). Model weights are prepared once before use (top). During use, all steps from image input to display and recording run on the personal device without network access (middle); the weight of the food is optional and, when entered, is appended to the fixed prompt as one sentence. Accuracy was evaluated in batch runs on reference image sets, and agreement between the application and the batch configuration was confirmed by an acceptance test (bottom). Only the research-protocol mode evaluated in this study is shown.

### 2.2. Image sets and reference values

NutriImage (NI887) comprises 887 photographs of dishes served at a university cafeteria in Kyoto, Japan (2002–2005), cropped to the dish to remove burned-in text [1,3]. Reference weight, energy and nutrients were calculated by registered dietitians from the ingredient-level recipe of each dish using the Standard Tables of Food Composition in Japan (Fifth Revised Edition, 2000). The median reference weight was 130 g and the median energy 168 kcal; images were 161–640 px wide (median 403 × 303 px). SNAPMe (SN1134) is a public set of photographs of free-living meals taken by US study participants before eating; the dataset authors linked each photograph to the food record that the participant entered in ASA24 (Automated Self-Administered 24-h Dietary Assessment Tool), and the reference values are taken from these records [4]. The records were not weighed, and whether the recorded portion equals the amount visible in the photograph was not verified for individual images. As in ref. [1], we used the 1,134 of the 1,272 before-meal images of non-packaged foods whose native resolution was 800 × 800 px, so that no image was resized; energy, protein, fat and carbohydrate are the ASA24 values, and salt equivalent was derived from sodium (Na mg × 2.54 / 1,000). Images with a reference energy of 0 kcal were excluded, leaving 886 and 1,131 images (883– 886 and 1,127–1,131 with a parsed estimate, depending on the model; Table 1). Further details are given in Online Resource 1.

**Table 1.**
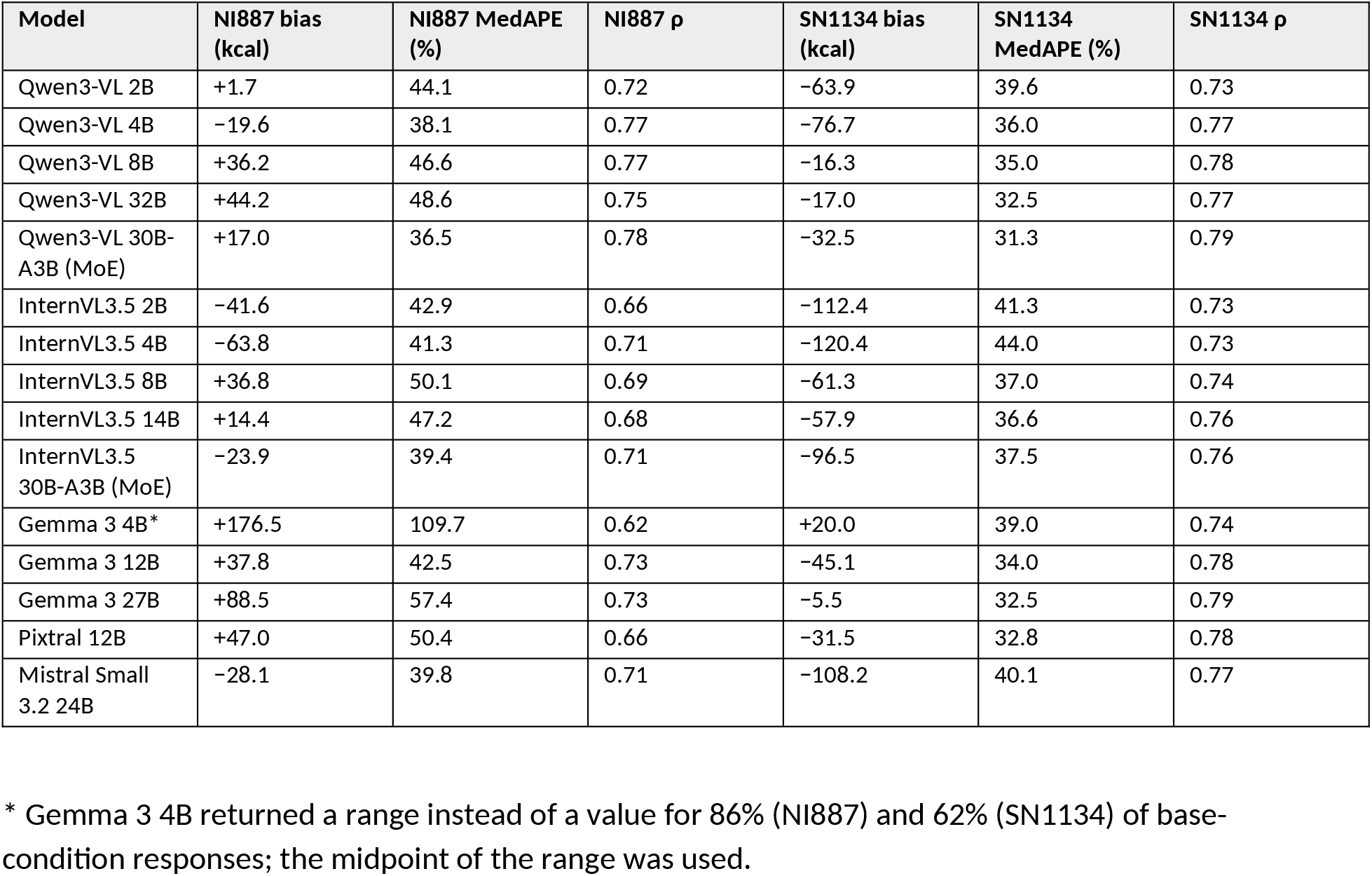
Base-condition accuracy of open vision–language models on a server (BF16, H100; unified reference tables). NI887: n = 886 (883–885 for three cells); SN1134: n = 1,127–1,131.

### 2.3. Prompt

The base prompt (Appendix A) was carried over verbatim from ref. [1]: it asks for energy, protein, fat, carbohydrate and salt from visual appearance only, without reading text in the image, in JSON with a one-line basis; the system message was empty. In the weight condition one sentence was appended: “The total weight of the food shown is {W} g.”, where W is the reference weight in grams.

### 2.4. Server-side comparison of open models (Stage 1)

Open models were run on one NVIDIA H100 (80 GB) of a cloud GPU service (Sakura Internet DOK) using vLLM 0.11 [5], in BF16 unless stated: Qwen3-VL-Instruct 2B, 4B, 8B, 30B-A3B (mixture of experts) and 32B [6]; Gemma 3 4B, 12B and 27B [7]; InternVL3.5 2B, 4B, 8B, 14B and 30B-A3B [8]; Pixtral-12B [9]; and Mistral Small 3.2 24B [10]. Official FP8 checkpoints of Qwen3-VL were also run. Generation used temperature 0 and at most 256 new tokens. Values were extracted from the JSON response with the same rules in all runs, including repair of malformed JSON; the handling of responses truncated at the token limit is described in Online Resource 1.

### 2.5. Local runs (Stage 2)

Desktop computer. Qwen3-VL-Instruct 2B, 4B and 8B quantized to 4 bits (MLX format [11]) were run on a Mac mini (2024; Apple M4, 24 GB unified memory) with mlx-vlm 0.6.17 [12], using the same prompt, images and generation settings.

Tablet. The 4-bit 4B weights were run on an iPad Air (M4, 12 GB, iPadOS 26) with a batch application built on the MLX Swift libraries [13] (mlx-swift 0.31.6; mlx-swift-lm commit 604fae7). The application loads the model and images only from local storage, makes no network request, and saves the raw model text, which was parsed with the same extraction code. A preliminary run of 30 images with an earlier library release (mlx-swift-lm 3.31.3), which contained an image-preprocessing defect, is reported separately.

Agreement between platforms was quantified as the proportion of images with an identical value and the mean paired difference.

Interactive interface. An application sharing the same inference path was implemented for iPhone and iPad (PLANS-iOS; Fig. 2). The user takes or selects a photograph and obtains the five values from the on-device model, may enter the food weight for re-estimation, and every request is stored locally without network access. The application resizes photographs to 1,024 px on the long side. As an acceptance test, 26 photographs of different meals (home-cooked, restaurant and packaged meals, taken by one of the authors) were analysed once each with the application on an iPhone 17 Pro (iOS 26.6.1), and the resized inputs saved by the application were processed by the batch application on the same iPhone under the base condition. The application also ran in airplane mode with Wi-Fi off (Fig. 2); accuracy was evaluated on the tablet only.

**Fig 2.**
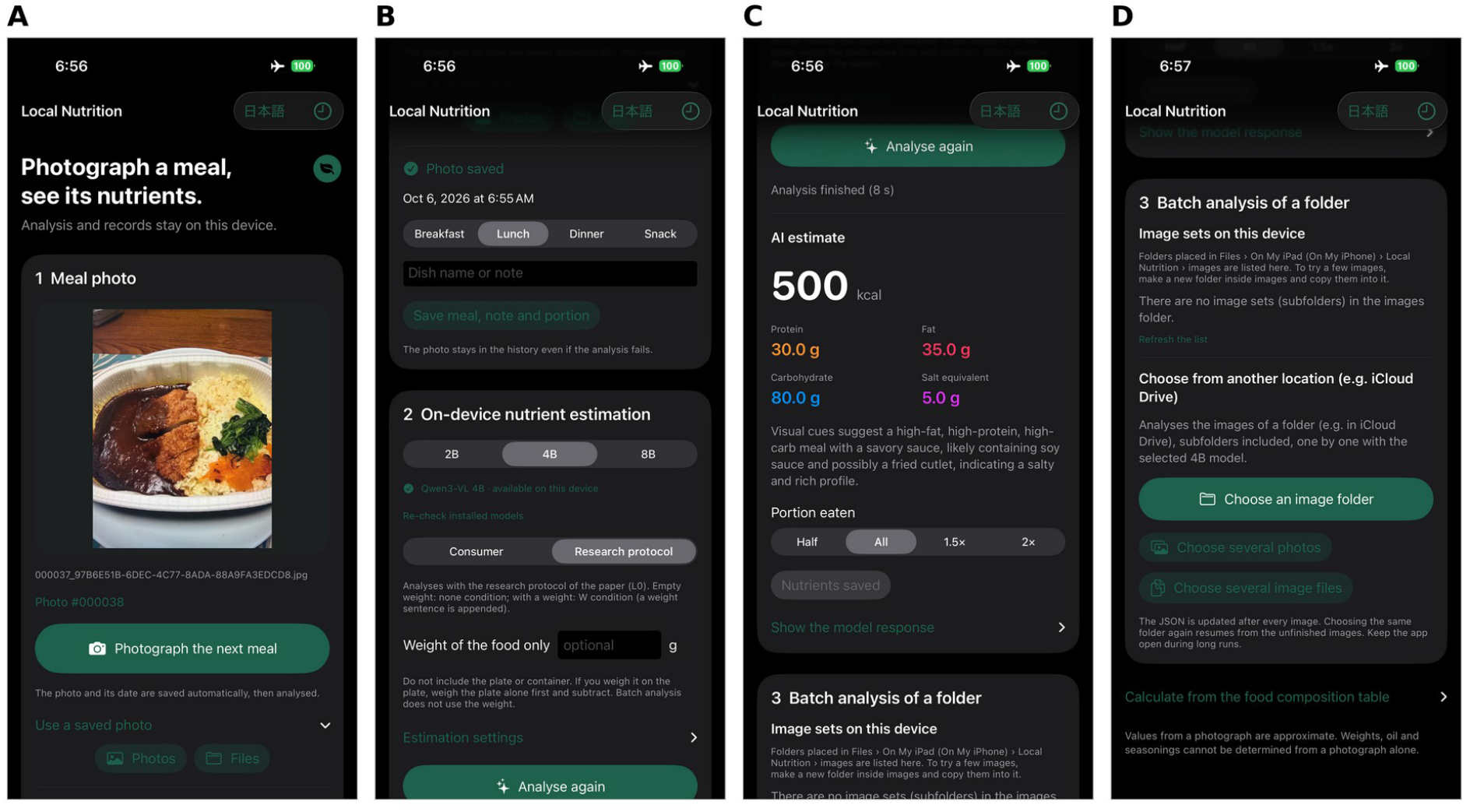
Screens of the interactive application (English display) on an iPhone 17 Pro in airplane mode with Wi-Fi off; Qwen3-VL-4B (4-bit), research-protocol mode. (A) Photographing a meal. (B) Model selection (2B/4B/8B), research protocol, and the optional field for the weight of the food alone, with the instruction to exclude the container. (C) Result of one estimate (8 s). Five values are displayed; their accuracy is reported in Section 3.4. The meal is a commercially available frozen product (labelled 615 kcal and 350 g; estimated 500 kcal); the screen illustrates the interface and is not an accuracy result. (D) Batch analysis of an image folder on the device.

### 2.6. Weight information and declared-weight error (Stage 3)

The effect of the food weight was evaluated on NI887, in which the weight of every dish is known, by comparing the base condition with the true weight (W × 1.0) for 2B, 4B and 8B on the desktop computer. To examine errors that a user might make, the 4B model was also run with the declared weight set to W × 0.5, 0.75, 0.9, 1.1, 1.25, 1.5 and 2.0 (multiplicative error), W + 150, 300 and 450 g (container weight not subtracted) and W rounded to the nearest 50 g (minimum 50 g), giving 9,757 new inferences. The desktop computer holds the same weights as the tablet, and all comparisons were between conditions on the same platform. Two quantities were defined in advance: the range of multiplicative error within which MedAPE was not worse than the base condition, and the tracking slope, the coefficient of the regression through the origin of log(estimate ÷ estimate at W × 1.0) on log(declared W ÷ true W), which is 1 when the estimate scales with the declared weight and 0 when the weight is ignored.

### 2.7. Reference runs of commercial models (Stage 4)

Six current commercial models (Claude Haiku 4.5 and Claude Sonnet 5, Anthropic; GPT-5.6 Sol and GPT-5.6 Luna, OpenAI; Gemini 3.8 Flash and Gemini 3.1 Pro, Google) were run through their APIs on the same images and prompt, with the vendor’s default temperature, one repetition and the lowest reasoning setting accepted (Online Resource 1, Table S1). Gemini 3.8 Flash was also run under the weight condition on NI887, with a second base-condition run for a paired comparison. These runs serve as a reference rather than a head-to-head benchmark, because temperature and image handling could not be made identical across vendors.

### 2.8. Outcome measures and statistics

Energy was the primary outcome; protein, fat, carbohydrate and salt equivalent were secondary. Accuracy was summarised by bias (mean of estimate minus reference), median absolute error (MedAE), median absolute percentage error (MedAPE), the proportion of images within ±20% of the reference, Spearman’s ρ and 95% limits of agreement. Images with a reference value of 0 for a nutrient were excluded from all accuracy measures of that nutrient (e.g., 51 NI887 images for salt; percentage error is undefined for them, and their estimates are reported in Online Resource 1). 95% confidence intervals (CIs) for MedAPE and for paired differences in MedAPE were obtained by bootstrap over images (2,000 resamples). Analyses used Python 3 (numpy, scipy).

## 3. Results

### 3.1. Selection of the model family and size

Table 1 shows base-condition accuracy on the server. In Qwen3-VL, ρ on NI887 rose from 0.72 (2B) to 0.77 (4B) and did not increase with the larger dense models, whose MedAPE was higher (8B 46.6% and 32B 48.6% vs 4B 38.1%) with overestimation (bias +36 and +44 kcal). The mixture-of-experts 30B-A3B gave the lowest MedAPE (36.5% and 31.3%) but would exceed the 12 GB memory of the tablet even at 4 bits. InternVL3.5 and Gemma 3 gave larger errors or lower ρ than Qwen3-VL-4B on NI887. FP8 checkpoints gave MedAPE within 1 point of BF16 (Table S2). Qwen3-VL-4B was selected for on-device use (4-bit weights 2.9 GB), with 8B as a comparator.

### 3.2. Effect of 4-bit quantization

On the desktop computer at 4 bits, the aggregate measures were close to those of the BF16 server runs for both 4B and 8B (Table 2; 4B: MedAPE 37.0% vs 38.1% on NI887 and 36.1% vs 36.0% on SN1134; paired difference on the same images −1.1 points, 95% CI −3.1 to +1.3, and +0.1 points, CI

**Table 2.** Base-condition accuracy of 4-bit (desktop computer, MLX) versus BF16 (server, vLLM) runs of Qwen3-VL on the unified reference tables (NI887 n = 886; SN1134 n = 1,131). The two configurations differ in precision, hardware, library and image preprocessing. ΔMedAPE: 4-bit minus BF16 on the same images, with 95% CI by paired bootstrap (2,000 resamples); identical: proportion of images with the same energy value in both runs.

| Model / set | Precision | Bias (kcal) | MedAPE (%) | p |
| --- | --- | --- | --- | --- |
| 4B, NI887 | BF16 | -19.6 | 38.1 | 0.77 |
|  | 4-bit | +1.1 | 37.0 | 0.76 |
| 4B, SN1134 | BF16 | -76.7 | 36.0 | 0.77 |
|  | 4-bit | -52.0 | 36.1 | 0.78 |
| 8B, NI887 | BF16 | +36.2 | 46.6 | 0.77 |
|  | 4-bit | +39.0 | 46.5 | 0.78 |
| 8B, SN1134 | BF16 | -16.3 | 35.0 | 0.78 |
|  | 4-bit | +0.6 | 36.2 | 0.78 |

ΔMedAPE (4-bit - BF16) [95% CI] and identical values: 4B NI887 -1.1 [-3.1, +1.3], 41%; 4B SN1134 +0.1 [-2.1, +1.8], 38%; 8B NI887 -0.1 [-2.3, +2.1], 77%; 8B SN1134 +1.2 [-0.2, +2.9], 67%.

−2.1 to +1.8), although the individual estimates were identical for only 38–41% of images (4B). Bias was shifted upward in the 4-bit runs (4B: +1.1 vs −19.6 kcal on NI887, −52.0 vs −76.7 kcal on SN1134). Because precision, hardware, library and image preprocessing all differed between the two configurations, the differences cannot be attributed to quantization alone. The response of 4B to the persona conditions of ref. [1] was retained in the 4-bit configuration (Japanese-minus-US dietitian −8.1 vs −13.1 kcal on NI887; −11.9 vs −12.6 kcal on SN1134), whereas for 8B it largely disappeared (−3.2 vs −15.4 and +1.0 vs −4.9 kcal; Online Resource 1, Table S3).

### 3.3. Reproduction on the tablet

On the tablet, 2,661 inferences on NI887 and 3,402 on SN1134 (three prompt conditions × 887 and 1,134 images) were completed without error; the base-condition analyses use the 886 and 1,131 images with a reference energy above 0 kcal. Base-condition accuracy was the same as on the desktop computer (Table 3). Energy values were identical to those on the desktop computer for 84% of NI887 images (mean paired difference +0.6 kcal; 78 higher, 67 lower) and 86% of SN1134 images (−1.6 kcal; 62 higher, 91 lower), and the other four values for 75–87% (Online Resource 1, Table S4). Among the images whose energy value differed, the absolute difference was mostly 50 kcal (maximum 120 kcal) and exceeded 50 kcal for 4% of all images (Online Resource 1, Table S5). In the acceptance test on the iPhone, the raw response (hence all five values) of the interactive application was identical to those of the batch application for all 26 photographs (median 7.9 s per image in the application, range 5–77 s, and 8.4 s in the batch application).

**Table 3.** Base-condition accuracy of Qwen3-VL-4B (4-bit) on the tablet and agreement with the desktop-computer run.

| Set | n | Bias (kcal) | MedAPE (%) | p | Identical energy vs desktop computer (%) | Mean difference vs desktop computer (kcal) | Time per image (s) |
| --- | --- | --- | --- | --- | --- | --- | --- |
| NI887 | 886 | +1.7 | 37.0 | 0.76 | 84 | +0.6 | 5.2 (9.8 over 8 h) |
| SN1134 | 1,131 | −53.5 | 36.5 | 0.78 | 86 | −1.6 | 12.6 |

With the earlier library release (3.31.3), 30 NI887 images gave lower values than the desktop computer for 61 of 90 image–condition pairs and higher values for none (mean −58 kcal; identical 32%); after the update, the same images agreed for 76% of pairs (mean +2.7 kcal). Processing time on the tablet was 5.2 s per NI887 image in a 30-image run and 9.8 s on average over the full 8-hour batch (the cause of the slowdown, such as thermal throttling, was not examined); SN1134 images (800 px) took 12.6 s. On the desktop computer the median was 3.2 s (4B, NI887). The 8B weights also ran on the tablet (9–19 s per image in a preliminary run).

### 3.4. Accuracy for the five values

Energy had the smallest relative error of the five values on the tablet (Table 4, Fig. 3). MedAPE for protein was 47.0% (NI887) and 44.4% (SN1134), for fat 61.1% and 61.4%, for carbohydrate 65.1% and 42.6%, and for salt 101.0% and 67.8%; ρ ranged from 0.51 (salt, NI887) to 0.85 (protein, SN1134).

**Table 4.**
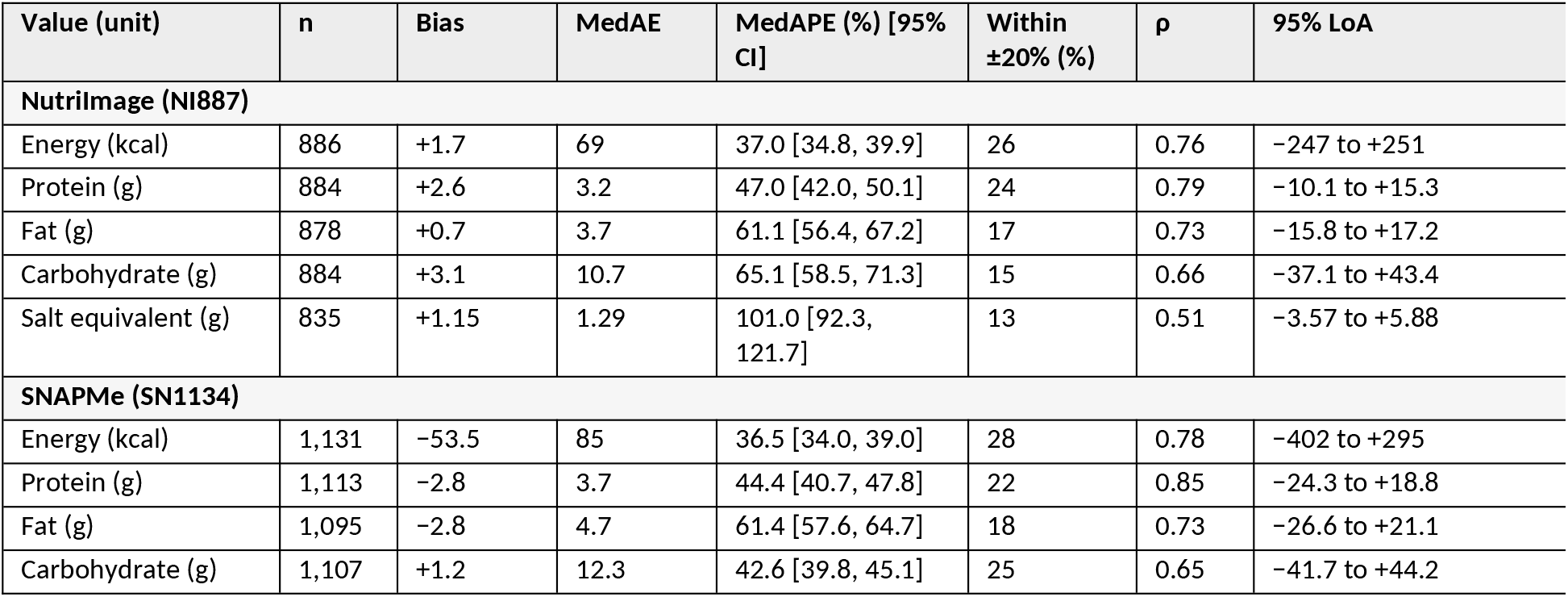
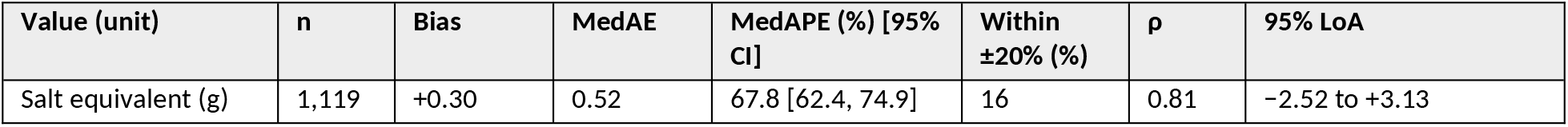
Accuracy of the five values estimated on the tablet (Qwen3-VL-4B, 4-bit, iPad Air; base condition). Bias, median absolute error (MedAE) and 95% limits of agreement (LoA, mean difference ± 1.96 SD) are in the unit of each value. Images with a reference value of 0 for the value concerned were excluded, hence the different n. Two NI887 dishes have reference salt values far above the others (24.7 g and 47.4 g; noted in the NutriImage README); excluding them, salt bias was +1.23 g and the LoA −2.05 to +4.51 g (n = 833).

**Fig 3.**
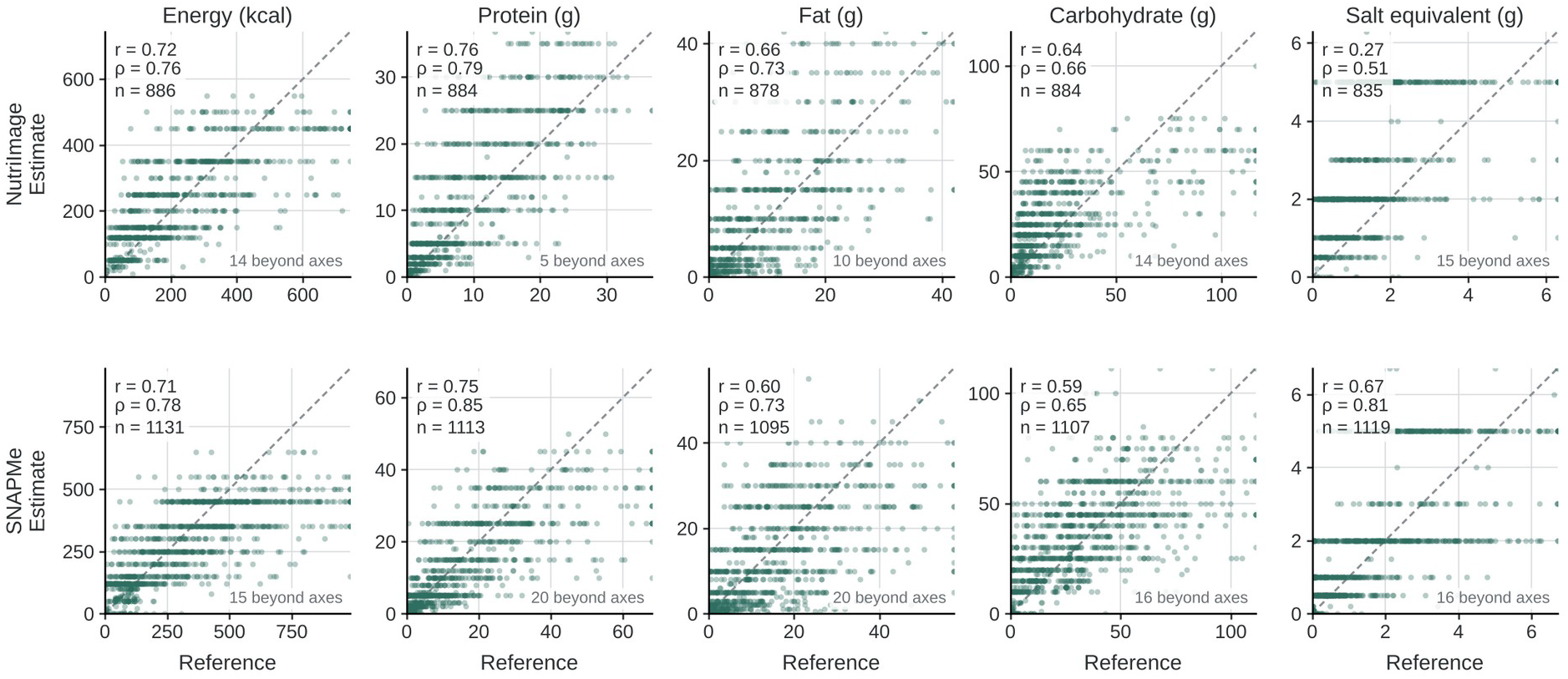
Estimated versus reference values of the five values on the tablet (Qwen3-VL-4B, 4-bit, base condition); top, NutriImage (NI887); bottom, SNAPMe (SN1134). Dashed line: identity. Bland–Altman plots for energy are given in Online Resource 1, Fig. S2. r, Pearson’s correlation; ρ, Spearman’s correlation. Axes are truncated at the 99th percentile of the pooled values; points beyond the axes are drawn at the edge and their number is shown, and all points are used for the correlations.

On NI887, salt was overestimated (bias +1.15 g): 41% of the dishes had a reference salt content below 1 g, whereas 74% of the estimates were 2–5 g. Estimates tended to take rounded values (e.g., 250 kcal or 2 g of salt), and the energy of large meals was underestimated: in the highest quintile of reference energy the median difference was −129 kcal (NI887) and −261 kcal (SN1134), and the lowest quintile was overestimated (+56 and +52 kcal; Online Resource 1, Fig. S2). The 95% limits of agreement for energy were −247 to +251 kcal (NI887) and −402 to +295 kcal (SN1134).

The ranking of model sizes differed by value (Online Resource 1, Table S6): on NI887, 2B gave a lower MedAPE than 4B for protein (41.9% vs 46.0%) and salt (45.9% vs 100.0%).

### 3.5. Weight information and errors in the declared weight

Supplying the true weight improved the 4B model on NI887 (Table 5): MedAPE fell from 37.0% to 32.5% (difference −4.5 points, 95% CI −7.3 to −2.1) and ρ rose from 0.76 to 0.84. The correction worked in both directions (−11.1 kcal on average for the 501 images overestimated without the weight, +34.6 kcal for the 385 underestimated). The 2B model improved similarly (MedAPE 43.0% to 38.3%), whereas for 8B ρ improved (0.78 to 0.82) but MedAPE did not (46.5% to 48.2%) and overestimation increased (bias +39 to +64 kcal). For the other values (4B), MedAPE fell most markedly for salt (100.0% to 66.8%; −33.2 points, CI −43.0 to −23.6) and marginally for protein (−4.2 points, CI −8.1 to −0.1; exploratory), with no clear change for fat (61.5% to 63.3%) or carbohydrate; for 2B, salt became worse (45.9% to 51.7%; +5.8 points, CI +1.6 to +10.5) (Online Resource 1, Table S7).

**Table 5.**
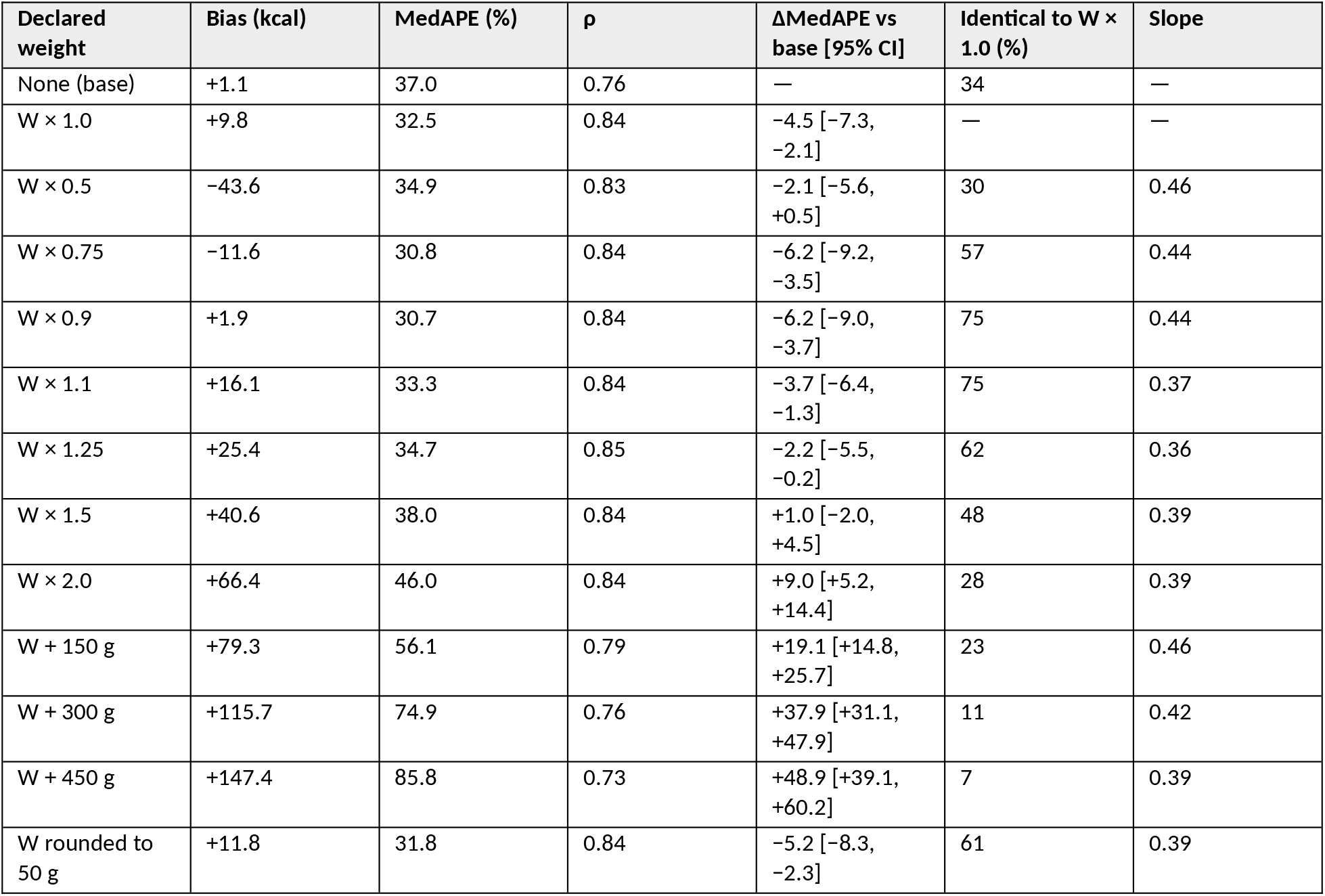
Accuracy of Qwen3-VL-4B (4-bit, desktop computer) on NI887 (n = 886) with the declared food weight varied. Slope: tracking slope of log(estimate) on log(declared weight), relative to the W × 1.0 run.

The estimate followed the declared weight only partially (Fig. 4): the pooled tracking slope over the multiplicative conditions was 0.42 (bootstrap 0.40–0.44), so doubling the declared weight raised the estimate about 1.3-fold. The point estimate of MedAPE was below that of the base condition for declared weights from 0.5× to 1.25× the true weight (the 95% CI of the difference excluded zero at 0.75×, 0.9×, 1.1× and 1.25×, and included it at 0.5×), was above it at 1.5× (38.0% vs 37.0%; +1.0 points, CI −2.0 to +4.5) and was clearly worse at 2.0× (46.0%; +9.0 points, CI +5.2 to +14.4). Rounding to 50 g gave MedAPE 31.8%. Adding 150, 300 or 450 g raised MedAPE to 56.1%, 74.9% and 85.8% and bias to +79, +116 and +147 kcal, and ρ fell to 0.73–0.79, whereas it remained 0.83–0.85 under multiplicative errors; for a dish of the median weight (130 g), however, +150 g corresponds to a declared weight of more than twice the true value, so the additive and multiplicative conditions are not matched in the size of the error.

**Fig 4.**
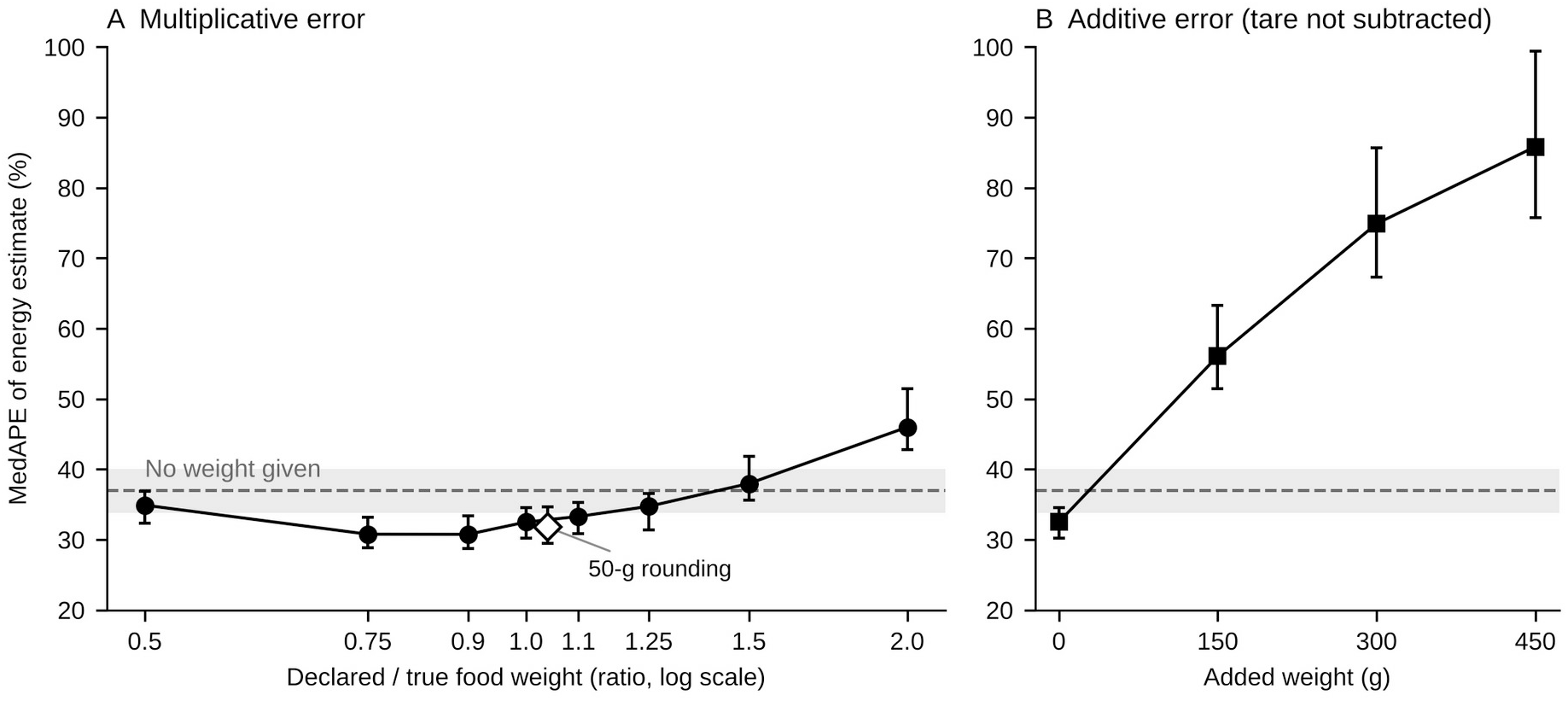
Median absolute percentage error of the energy estimate (Qwen3-VL-4B, 4-bit; NI887, n = 886) as a function of the declared food weight. (A) Multiplicative error (declared ÷ true weight, log scale); the open diamond is the weight rounded to the nearest 50 g, plotted just to the right of 1.0 to avoid overlap (the declared ÷ true ratio of this condition varies by image: median 1.01, 5th–95th percentiles 0.76–1.32). (B) Additive error (grams added to the true weight, as when the container is not subtracted). Dashed line and grey band: MedAPE without weight information and its 95% CI. Error bars: 95% CI by paired bootstrap over images.

The lowest MedAPE was at 0.75× and 0.9× the true weight (30.8% and 30.7%), possibly because under-declaration offset the positive bias of the 4-bit run; we do not regard this as a property to be exploited.

### 3.6. Reference comparison with commercial models

On the same images (energy only; Table 6), the best commercial model was Gemini 3.8 Flash (MedAPE 28.5% on NI887 and 28.9% on SN1134); the others ranged from 34.7% to 97.2% on NI887 and from 30.1% to 50.3% on SN1134. The on-device 4B model (37.0% and 36.5%) was within this range, about 8 points behind Gemini 3.8 Flash and, on NI887, close to Claude Sonnet 5 and Gemini 3.1 Pro. Supplying the true weight improved Gemini 3.8 Flash by a similar amount (NI887 MedAPE from 28.4% in the paired base-condition run to 22.9%; −5.5 points, CI −7.9 to −3.1; ρ 0.86 to 0.92), so the gap was not changed by weight information.

**Table 6.** Reference comparison of operating conditions and base-condition accuracy (unified reference tables). Commercial models: vendor default temperature, one repetition, lowest reasoning setting accepted.

| Configuration | Images processed | Network | NI887 MedAPE (%) / $p$ | NI887 with weight MedAPE (%) / $p$ | SN1134 MedAPE (%) / $p$ |
| --- | --- | --- | --- | --- | --- |
| Qwen3-VL-4B 4-bit, iPad Air (M4) | On device | None | 37.0 / 0.76 | — | 36.5 / 0.78 |
| Qwen3-VL-4B 4-bit, Mac mini (M4) | On device | None | 37.0 / 0.76 | 32.5 / 0.84 | 36.1 / 0.78 |
| Qwen3-VL-4B BF16, H100 server | Cloud GPU (benchmark run)* | Internet* | 38.1 / 0.77 | — | 36.0 / 0.77 |
| Gemini 3.8 Flash | Vendor cloud | Internet | 28.5 / 0.86 | 22.9 / 0.92 | 28.9 / 0.82 |
| Gemini 3.1 Pro | Vendor cloud | Internet | 34.7 / 0.80 | — | 30.7 / 0.81 |
| Claude Sonnet 5 | Vendor cloud | Internet | 40.8 / 0.79 | — | 30.1 / 0.80 |
| Claude Haiku 4.5 | Vendor cloud | Internet | 50.4 / 0.60 | — | 35.6 / 0.72 |
| GPT-5.6 Sol | Vendor cloud | Internet | 51.3 / 0.77 | — | 33.2 / 0.80 |
| GPT-5.6 Luna | Vendor cloud | Internet | 97.2 / 0.72 | — | 50.3 / 0.79 |

\* The server runs were benchmark runs on a cloud GPU service (Section 2.4). The configuration is intended for deployment on an institutional server reached over the local network, which would keep photographs within the institution; such a deployment was not tested in this study.

## 4. Discussion

### 4.1. Principal findings

A 4B open VLM quantized to 4 bits gave, on a consumer tablet without network access, energy-estimation accuracy whose aggregate measures were close to those of the BF16 model on a cloud GPU, within the range of current commercial models although below the best of them; the other four values were estimated less accurately. Within the configurations evaluated, model size beyond 4B did not improve accuracy on NI887, and the change of precision and execution stack altered the aggregate measures little; the food weight supplied as text improved the energy and salt estimates of the 4B model on NI887 and tolerated proportional errors of up to about 25% and rounding to 50 g but not inclusion of the container weight; and the version of the inference library changed the estimate by tens of kcal.

### 4.2. Implications for design

Model choice. On NI887, accuracy did not improve beyond 4B and the larger dense Qwen3-VL models overestimated, whereas on SN1134 MedAPE decreased slightly with size (36.0% at 4B, 32.5% at 32B and 31.3% at 30B-A3B; Table 1). Whether the ceiling for a small model is set by dish identification rather than by scale [1] was not tested, because identification and portion estimation were not evaluated separately. The 8B model was also less stable in the 4-bit configuration in its response to prompt content and did not benefit from weight information. We therefore regard 4B as the best compromise among the evaluated configurations for a 12-GB tablet rather than as an optimum; the ranking of sizes differed for the other values (Online Resource 1, Table S6). Model choice for a device should rest on evaluation under the deployment protocol rather than on parameter count.

Weight entry. A tracking slope of about 0.4 shows that the estimate responded to the declared weight only partially; the mechanism was not examined. Within the conditions examined, the energy estimate stayed within the base-condition error for proportional errors of up to about 25% and for rounding to 50 g, so an approximate weight may still be useful. The error that clearly degraded accuracy was the inclusion of the container weight (150 g, roughly a small bowl, raised MedAPE by 19 points). The interface therefore asks for the weight of the food alone and instructs the user to subtract the container (Fig. 2B).

Acceptance test. A defect in one library release shifted the estimate by −58 kcal on average without any change to the model, which would not be detected by an accuracy figure from a server evaluation. We suggest that an on-device deployment be accepted only after it reproduces, on a fixed image set, the outputs of the evaluated configuration; the proportion of identical outputs and the distribution of the remaining differences are practical quantities (here 84–86% identical and 96% of images within 50 kcal); the acceptance threshold should be set from the difference tolerable for the intended use. The acceptance test of the interactive application covered only the image path.

### 4.3. Clinical context

PLANS was motivated by medical institutions that use meal photographs in guidance for diabetes and dyslipidaemia but cannot send them outside. A MedAPE of about 35% means that half of the estimates fall outside roughly ±35% of the reference; about a quarter were within ±20%, and the limits of agreement were wide for all values (Table 4), fat and salt in particular (MedAPE about 61% and 68–101%). The estimates are therefore a candidate input for counselling whose usefulness has not been tested; evaluation at the level of patients and of several meals is required. Human estimation from images is also imprecise [14–16], but the tasks and measures differ from ours, and we do not regard the present results as evidence that the model replaces a dietitian’s assessment. In repeated runs of the same configuration the same input gave the same output, so the effect of a single change—in how a meal is photographed or in the prompt—can be examined in isolation; comparing estimates for packaged foods served on a plate with the values stated on the package may provide one benchmark for adapting the model. Beyond dietary assessment, the same arrangement—an image captured on the device, a fixed prompt, structured output and local storage —applies to other image-based tasks in institutions that restrict transmission to cloud AI services, such as reading forms or checking labels. Each task would require its own evaluation; the present implementation and acceptance procedure may serve as a starting point.

### 4.4 Relation to previous work

Offline calorie-tracking applications using a small VLM have been released [2]; the present study differs in evaluating design factors against reference values and in reporting agreement between platforms. In Japan, an AI-based dietary recording application showed moderate agreement with weighed records, with overestimation of energy [17]. Evaluations of quantized models on mobile devices have mainly reported processing time, memory and energy use [18,19]; we add the agreement of the estimates themselves.

### 4.5 Limitations

The study used one model generation and one execution stack (MLX on Apple silicon); other hardware and libraries may differ. Device data came from one iPad Air; accuracy on the iPhone was not evaluated. Reference values are recipe-based (NI887, from the Fifth Revised Edition of the Standard Tables, which differs from the current Eighth Revised Edition in energy conversion and dietary fibre) or participant-recorded food records (SN1134, ASA24), not weighed intake. Percentage errors for small reference amounts, notably salt, are sensitive to small absolute differences; the commercial comparison was limited to energy. The 4-bit runs had a positive offset of bias (up to about 25 kcal) whose mechanism was not examined, so absolute bias should be taken from the deployed platform. Model selection and evaluation used the same two image sets, which may favour a configuration that performs well on them. The application uses 1,024-px photographs, whereas the evaluated images were smaller, and accuracy at 1,024 px was not evaluated. The declared-weight experiment used one model and one image set. The server and local runs differed in one parsing rule: responses truncated at the token limit without a closing brace were treated as failures on the server but accepted locally when all five values were present; this concerned only the 2B model (at most 88 responses per set) and hence the size comparison, not the 4B results. SNAPMe contains several images per participant, and the image-level bootstrap does not account for this clustering. Photographs taken by patients in free-living conditions, the weight-entry path of the interface and its usability were not examined, and the estimates were not validated on images independent of those used for model selection.

## 5. Conclusions

A 4B VLM quantized to 4 bits gave, on a consumer tablet without network access, energy-estimation accuracy whose aggregate measures were close to those of the server-side BF16 run; protein, fat, carbohydrate and salt were estimated less accurately. Within the configurations evaluated, model size and precision changed the aggregate measures little; the food weight, when supplied, improved the energy and salt estimates of the selected model, and the energy estimate tolerated proportional errors of up to about 25% and rounding to 50 g in the declared weight but not inclusion of the container weight; and the inference library version warranted an acceptance test before deployment. These observations may serve as design considerations for offline dietary tools in institutions where photographs cannot leave the premises; the usefulness of the estimates in counselling remains to be examined with photographs taken by users.

## Supporting information

Supplementary Information (Online Resource 1)

## Data Availability

All data produced in the present study are available online. Per-image model outputs, the analysis scripts, the acceptance-test records and 23 of the 26 acceptance-test photographs (three are withheld because personal items are visible) are available on Zenodo (https://doi.org/10.5281/zenodo.23298787). The source code of PLANS-iOS (v0.1.0-paper2, MIT License) is available at https://github.com/shnkgw-rincom/PLANS-iOS and archived at https://doi.org/10.5281/zenodo.23309553. The SNAPMe dataset is openly available in Ag Data Commons (https://doi.org/10.15482/USDA.ADC/1528346). The NutriImage reference table and README are openly available on Zenodo (https://doi.org/10.5281/zenodo.21961032); the 887 images are available for academic research under restricted access through the access-request function of the repository. The raw outputs of the preliminary run with mlx-swift-lm 3.31.3 are not deposited.

https://github.com/shnkgw-rincom/PLANS-iOS

https://doi.org/10.5281/zenodo.23309553

https://doi.org/10.15482/USDA.ADC/1528346

## Acknowledgments

None.

## Appendix

A. Prompt

Base prompt (user message; system message empty): Look at this food image. Estimate the nutritional content based solely on visual appearance—color, shape, ingredients, and portion size. Do NOT read any text, labels, or numbers visible in the image. Use only visual information. Provide a brief one-line explanation of your reasoning. Respond ONLY in JSON format: {energy_kcal: value, protein_g: value, fat_g: value, carb_g: value, salt_g: value, basis: one-line reasoning}

Weight condition: the sentence “The total weight of the food shown is {W} g.” is appended to the user message.

## Supplementary Information

Online Resource 1 (ESM_1.docx): Supplementary Methods; Fig. S1, study flow; Fig. S2, Bland–Altman plots for energy on the tablet; Table S1, settings of the commercial models; Table S2, BF16 and FP8 checkpoints of Qwen3-VL; Table S3, persona-condition contrasts; Table S4, agreement of the five values between tablet and desktop computer; Table S5, distribution of the differences between tablet and desktop computer; Table S6, accuracy of the five values by model size; Table S7, accuracy of the five values under the weight and declared-weight conditions.

## Declarations

### Ethics approval

The study used existing anonymized image data and reference values (NutriImage, curated by the authors; SNAPMe, a public dataset) and did not involve human participants; it was therefore judged by the authors not to require ethical review. The 26 acceptance-test photographs are meals photographed by one of the authors solely to test the application; no person is depicted and no personal data were collected. Three of these photographs show personal items in the background and are not deposited.

### Consent to participate

Not applicable.

### Consent for publication

Not applicable.

### Use of generative AI tools

Generative AI tools were used between September and October 2026 to assist with writing the analysis scripts and the application code, checking references, and drafting and editing the text: Claude (Anthropic; Claude Sonnet 4.6, Claude Opus 5.5 and Claude Fable 5.1), ChatGPT (OpenAI; GPT-6, Instant mode) and the Codex application (OpenAI; GPT-6-based model; used from mid-September 2026, most recent version 26.1007.21434), accessed through the vendors’ applications. The tools were instructed in conversation with the study’s files, scripts and draft text; no fixed prompt was used. All reported values were recomputed from the per-image output files with the analysis scripts and checked by the authors, who reviewed all output and take full responsibility for the content. The commercial models of Section 2.7 were study subjects, not writing aids.

### Data availability

The NutriImage dataset is archived on Zenodo (concept DOI 10.5281/zenodo.21944881; version 1.1 used in this study, 10.5281/zenodo.21961032); the reference table and a README documenting provenance are openly available, and the 887 text-removed images are available for academic research under restricted access via the repository’s access-request function (redistribution is not permitted). The SNAPMe dataset is openly available in Ag Data Commons (https://doi.org/10.15482/USDA.ADC/1528346) [4]. Per-image model outputs, including the acceptance-test records and 23 of the 26 acceptance-test photographs (three are withheld because personal items are visible), are available at https://doi.org/10.5281/zenodo.23298787. The raw outputs of the preliminary run with mlx-swift-lm 3.31.3 (Section 3.3) are not deposited.

### Code availability

The analysis scripts and the declared-weight input files are included in the data package above. The source code of PLANS-iOS (MIT License) and of the tablet batch application (tag paper2-ipad-run) is available at https://github.com/shnkgw-rincom/PLANS-iOS; the version evaluated in this study (v0.1.0-paper2) is archived at https://doi.org/10.5281/zenodo.23309553.

### Author contributions

Shinichi Nakagawa conceived and designed the study, developed the software, carried out the experiments and the analysis, prepared the figures, and wrote the first draft of the manuscript. Akira Yamamoto contributed to validation, supervised the study, and reviewed and edited the manuscript. Both authors read and approved the final manuscript.

