## Supplementary Information (Online Resource 1) for "Development and Evaluation of an Offline Vision–Language Model System for Estimating Nutrients from Meal Photographs on Personal Devices: The PLANS (Personal-device Local Assessment of Nutrients System) Study"

### Supplementary Methods

*Image sets*. NutriImage photographs were taken at a university co-op cafeteria in Kyoto under the "Health and Nutrition Information" project (2002–2005). The images depict prepared food only; no persons are shown. They are the authors' own and have been curated by the corresponding author since the original project. Reference values were calculated by registered dietitians from the ingredient-level recipe of each served dish together with the total served weight (foodamt_g). One item (oolong tea, 0 kcal) was excluded, leaving 886 images. Image sizes ranged from 161 to 640 px in width and from 121 to 480 px in height. The reference table and a README documenting provenance are openly available on Zenodo; the text-removed images are available for academic research under restricted access via the repository's access-request function. SNAPMe [4] contains 1,475 before-meal photographs of non-packaged foods taken by 95 US participants, each linked by the dataset authors to the participant's ASA24 food record for that eating occasion; the records were entered by the participants and quality-controlled by the study staff, and no weighing or dietitian re-assessment was involved. Of the 1,272 before-meal images of the previous study [1], the 1,134 whose native resolution was exactly 800 × 800 px (JPEG) were used; the remaining 138 higher-resolution images were excluded to keep the input resolution uniform without resizing (reference energy of retained and excluded images: mean 313 vs 319 kcal). The reference values for energy, protein, fat and carbohydrate are the ASA24 values linked to each photograph (summed over the record lines linked to the same photograph); salt equivalent was derived from sodium as Na (mg) × 2.54 / 1,000. Because the record describes the portion the participant reported and the photograph shows the food before eating, the two may differ for individual images (e.g., food not eaten or added later); this was not verified image by image. Three images with a reference energy of 0 kcal were excluded, leaving 1,131. The same reference tables were used for all runs.

*Prompt*. In the weight condition, W was written with up to six significant digits (e.g., 67.5, 241.84). No other wording was varied.

*Server runs*. The server runs used a container on a cloud GPU service (Sakura Internet DOK), with a single image per request; they served to compare models, and no institutional server was used. Numeric values were extracted from the JSON response; when the JSON was malformed, keys were repaired or values were recovered by a regular expression, and when a range was returned its midpoint was used. In the server runs, responses without a closing brace were treated as failures, following ref. [1]. In the local runs (desktop computer and tablet), responses truncated at the 256-token limit were retained when all five values preceded the truncation, which occurred only in the repeated one-line basis of the 2B model (base condition: 20 of 887 NI887 and 88 of 1,134 SN1134 responses; weight condition: 64 of 887); no response of the 4B or 8B model lacked a closing brace, and no local response failed to parse.

*Local runs*. On the desktop computer, image preprocessing followed the model's preprocessor configuration (pixel limits as shipped with the model). The tablet batch application reads the model and image files from its Documents folder, applies the model's preprocessor pixel limits explicitly, uses an empty system message for the base condition, temperature 0 and 256 tokens, and writes the raw model text to a JSON-lines file. The defect of mlx-swift-lm 3.31.3 concerned the image colour transfer curve and was corrected on the main branch. The interactive application is displayed as "Local Nutrition" in the version shown in Fig. 2; on the iPhone 17 Pro the 4-bit 4B model took 5–11 s per image for 24 of the 26 acceptance-test photographs (median 7.9 s); two images took 30 s and 77 s, with identical outputs. A 27th photograph, a second shot of one of the 26 meals, also gave an identical raw response; it is excluded from the count of 26 and marked in the records. The acceptance-test records (application export, batch output and the paired comparison) are included in the data package.

*Commercial models.* Cost and processing time were not compared, because services that require photographs to be sent outside the institution cannot be used on the institutional terminals assumed here.

*Outcome measures.* MedAPE is the median of the absolute difference divided by the reference value. Pearson's r is shown in the scatter plots (Fig. 3 of the main text). Processing time per image was recorded by the application.

*Images with a reference value of 0.* For the five values, 0 in the reference table occurred for protein (NI887 2 images, SN1134 18), fat (8, 36), carbohydrate (2, 24) and salt (51, 12); none for energy after the exclusion described above. For these images the tablet (4B, base condition) returned 0 in most cases: protein 2/2 and 15/18, fat 8/8 and 34/36, carbohydrate 1/2 and 16/24, salt 30/51 and 5/12 (NI887 and SN1134). The remaining estimates were small (maximum 2.0 g of salt on NI887, 28 g of carbohydrate and 0.1 g of salt on SN1134). These images were excluded from all accuracy measures of the value concerned.

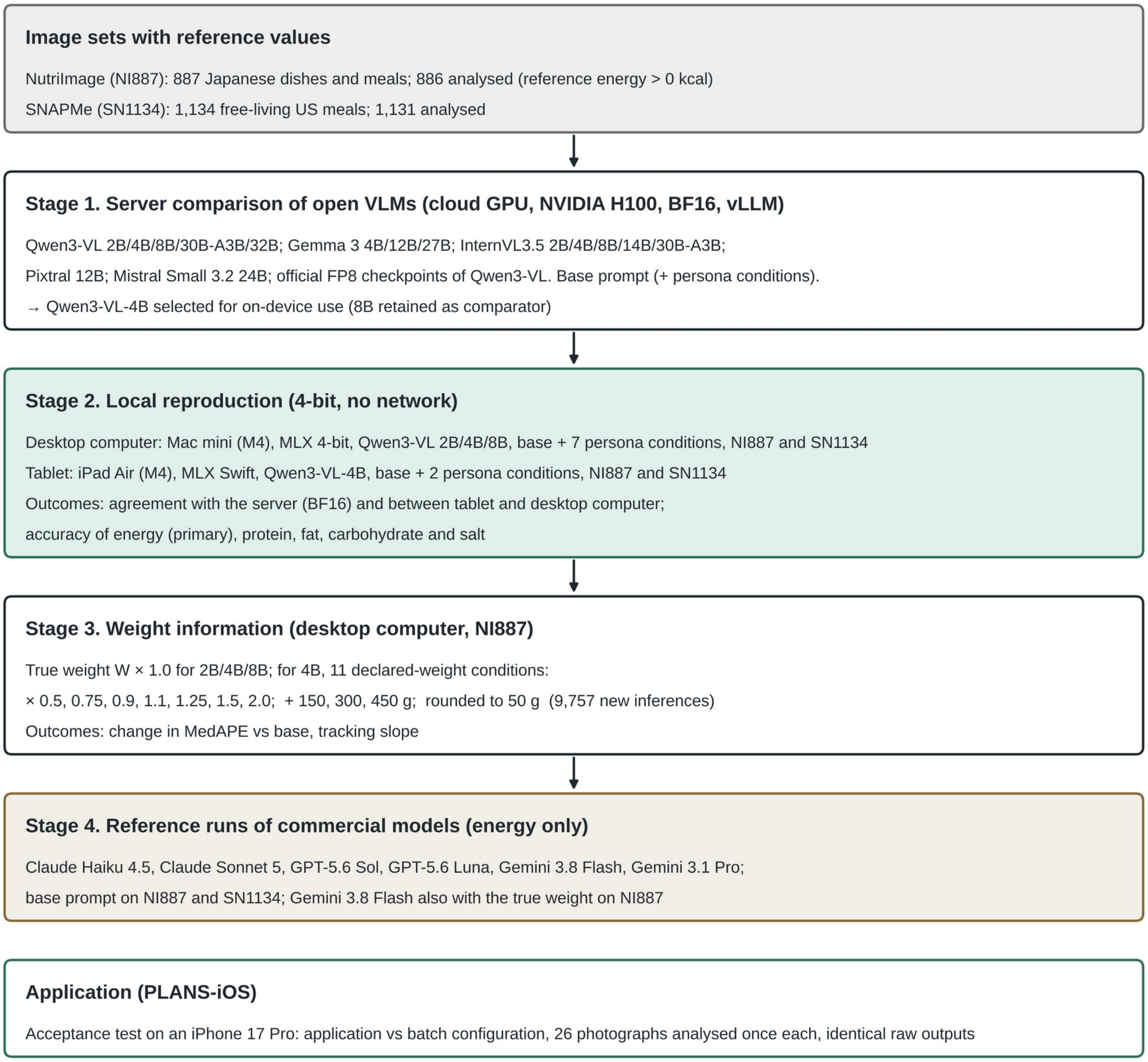


**Fig. S1. Study flow. Analysed numbers exclude images with a reference energy of 0 kcal. Persona conditions were included only to examine whether a model's response to prompt content was retained after quantization; they are not part of PLANS.**

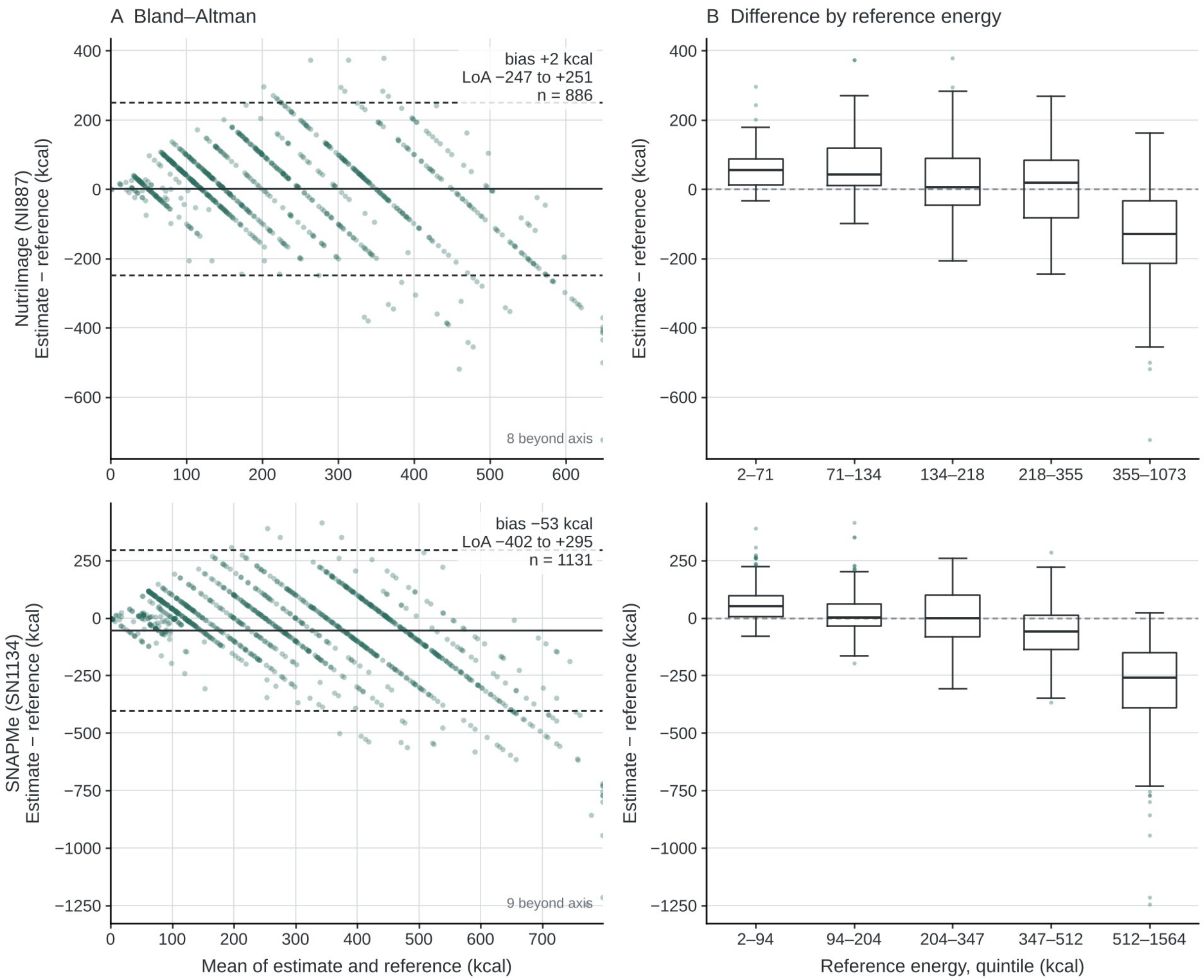


**Fig. S2. Agreement of the energy estimate on the tablet (Qwen3-VL-4B, 4-bit, base condition) with the reference. (A) Bland–Altman plots; solid line, mean difference; dashed lines, 95% limits of agreement (mean ± 1.96 SD). The diagonal streaks arise from estimates taking rounded values. (B) Difference by quintile of reference energy; boxes, median and interquartile range; whiskers, 1.5 × interquartile range; points, values beyond the whiskers.**

**Table S1. Settings of the commercial models (API).**

| Model | Vendor | Model identifier | Temperature | Reasoning / thinking setting | Run dates |
|---|---|---|---|---|---|
| Claude Haiku 4.5 | Anthropic | claude-haiku-4-5-20251001 | vendor default | thinking disabled | 2026-09-13 to 09-14 |
| Claude Sonnet 5 | Anthropic | claude-sonnet-5 | vendor default | thinking disabled | 2026-09-13 to 09-14 |
| GPT-5.6 Sol | OpenAI | gpt-5.6-sol | vendor default | reasoning effort none | 2026-09-13 to 09-14 |
| GPT-5.6 Luna | OpenAI | gpt-5.6-luna | vendor default | reasoning effort none | 2026-09-13 to 09-14 |
| Gemini 3.8 Flash | Google | gemini-3.8-flash | vendor default | thinking budget 0 | 2026-09-13 to 09-14; second base-condition run on NI887, |

| Model | Vendor | Model identifier | Temperature | Reasoning / thinking setting | Run dates |
|---|---|---|---|---|---|
| | | | | | 2026-09-22; weight condition on NI887, 2026-09-28 |
| Gemini 3.1 Pro | Google | gemini-3.1-pro-preview | vendor default | thinking level low | 2026-09-13 to 09-14 |

All runs used the same images and base prompt as the open models (Appendix A), with one repetition per image and the vendor's default sampling temperature. Maximum output tokens: 256 (Anthropic and OpenAI models) and 2,048 (Google models). Run dates are for NI887 and SN1134 unless stated.

**Table S2. Base-condition energy accuracy of Qwen3-VL in BF16 and in the official FP8 checkpoints (server, H100).**

| Model | Set | Precision | n | Bias (kcal) | MedAPE (%) | Identical to BF16 (%) |
|---|---|---|---|---|---|---|
| Qwen3-VL 2B | NI887 | BF16 | 885 | +1.7 | 44.1 | — |
| Qwen3-VL 2B | NI887 | FP8 | 885 | +10.4 | 44.7 | 83 |
| Qwen3-VL 2B | SN1134 | BF16 | 1,130 | −63.9 | 39.6 | — |
| Qwen3-VL 2B | SN1134 | FP8 | 1,130 | −56.7 | 39.1 | 82 |
| Qwen3-VL 4B | NI887 | BF16 | 886 | −19.6 | 38.1 | — |
| Qwen3-VL 4B | NI887 | FP8 | 886 | −16.5 | 37.4 | 86 |
| Qwen3-VL 4B | SN1134 | BF16 | 1,131 | −76.7 | 36.0 | — |
| Qwen3-VL 4B | SN1134 | FP8 | 1,131 | −77.6 | 35.9 | 89 |
| Qwen3-VL 8B | NI887 | BF16 | 886 | +36.2 | 46.6 | — |
| Qwen3-VL 8B | NI887 | FP8 | 886 | +36.2 | 46.9 | 89 |
| Qwen3-VL 8B | SN1134 | BF16 | 1,131 | −16.3 | 35.0 | — |
| Qwen3-VL 8B | SN1134 | FP8 | 1,131 | −16.1 | 35.3 | 87 |
| Qwen3-VL 30B-A3B | NI887 | BF16 | 886 | +17.0 | 36.5 | — |
| Qwen3-VL 30B-A3B | NI887 | FP8 | 886 | +18.0 | 36.9 | 80 |
| Qwen3-VL 30B-A3B | SN1134 | BF16 | 1,131 | −32.5 | 31.3 | — |
| Qwen3-VL 30B-A3B | SN1134 | FP8 | 1,131 | −32.8 | 31.2 | 78 |
| Qwen3-VL 32B | NI887 | BF16 | 886 | +44.2 | 48.6 | — |
| Qwen3-VL 32B | NI887 | FP8 | 886 | +42.6 | 47.7 | 91 |
| Qwen3-VL 32B | SN1134 | BF16 | 1,131 | −17.0 | 32.5 | — |
| Qwen3-VL 32B | SN1134 | FP8 | 1,131 | −16.1 | 32.6 | 92 |

Same images, prompt and settings as in Table 1 of the main text (temperature 0, 256 tokens). Identical to BF16: proportion of images whose energy estimate was the same in the FP8 and BF16 runs. Values are computed on the images parsed in both runs. For 2B, one NI887 response and one SN1134 response of the BF16 run lacked a closing brace and were treated as failures, as in Table 1 (n = 885 and 1,130); all FP8 responses were parsed.

**Table S3. Persona-condition contrasts in the energy estimate (paired by image; mean difference in kcal) at 4 bits on the desktop computer and tablet, and in BF16 on the server.**

| Set | Platform | Model | Contrast | n | 4-bit mean difference [95% CI] | Identical (%) | BF16 server mean difference |
|---|---|---|---|---|---|---|---|
| NI887 | Desktop computer (MLX 4-bit) | 2B | Japanese dietitian − US dietitian | 886 | +0.0 [−2.0, +2.0] | 74 | +2.8 |
| NI887 | Desktop computer (MLX 4-bit) | 2B | larger portion − smaller portion | 886 | +4.1 [+3.1, +5.2] | 86 | +3.6 |
| NI887 | Desktop computer (MLX 4-bit) | 2B | Japanese home − US home | 886 | −10.6 [−12.3, −8.9] | 70 | −3.8 |
| NI887 | Desktop computer (MLX 4-bit) | 4B | Japanese dietitian − US dietitian | 886 | −8.1 [−9.9, −6.0] | 74 | −13.1 |
| NI887 | Desktop computer (MLX 4-bit) | 4B | larger portion − smaller portion | 886 | +18.1 [+15.9, +20.2] | 69 | +21.9 |
| NI887 | Desktop computer (MLX 4-bit) | 4B | Japanese home − US home | 886 | −11.4 [−13.5, −9.3] | 71 | −18.1 |
| NI887 | Desktop computer (MLX 4-bit) | 8B | Japanese dietitian − US dietitian | 886 | −3.2 [−5.5, −1.0] | 72 | −15.4 |
| NI887 | Desktop computer (MLX 4-bit) | 8B | larger portion − smaller portion | 886 | +29.8 [+27.0, +32.8] | 56 | +44.2 |
| NI887 | Desktop computer (MLX 4-bit) | 8B | Japanese home − US home | 886 | −9.0 [−10.9, −7.1] | 75 | −17.2 |
| NI887 | Tablet (iPad, MLX Swift 4-bit) | 4B | Japanese dietitian − US dietitian | 886 | −9.2 [−11.1, −7.3] | 74 | −13.1 |
| SN1134 | Desktop computer (MLX 4-bit) | 2B | Japanese dietitian − US dietitian | 1131 | −2.5 [−5.0, −0.5] | 80 | +2.5 |
| SN1134 | Desktop computer (MLX 4-bit) | 2B | larger portion − smaller portion | 1131 | +1.6 [+0.7, +2.6] | 89 | +3.3 |
| SN1134 | Desktop computer (MLX 4-bit) | 2B | Japanese home − US home | 1131 | −11.1 [−13.7, −9.1] | 75 | −4.2 |
| SN1134 | Desktop computer (MLX 4-bit) | 4B | Japanese dietitian − US dietitian | 1131 | −11.9 [−13.7, −10.1] | 72 | −12.6 |
| SN1134 | Desktop computer (MLX 4-bit) | 4B | larger portion − smaller portion | 1131 | +15.7 [+13.9, +17.6] | 72 | +18.5 |
| SN1134 | Desktop | 4B | Japanese home | 1131 | −13.1 [−15.0, | 70 | −14.8 |

| Set | Platform | Model | Contrast | n | 4-bit mean difference [95% CI] | Identical (%) | BF16 server mean difference |
|---|---|---|---|---|---|---|---|
| | computer (MLX 4-bit) | | – US home | | −11.3] | | |
| SN1134 | Desktop computer (MLX 4-bit) | 8B | Japanese dietitian – US dietitian | 1131 | +1.0 [−1.3, +3.0] | 76 | −4.9 |
| SN1134 | Desktop computer (MLX 4-bit) | 8B | larger portion – smaller portion | 1131 | +20.4 [+18.2, +22.5] | 70 | +32.0 |
| SN1134 | Desktop computer (MLX 4-bit) | 8B | Japanese home – US home | 1131 | −5.5 [−7.2, −4.0] | 81 | −12.4 |
| SN1134 | Tablet (iPad, MLX Swift 4-bit) | 4B | Japanese dietitian – US dietitian | 1131 | −12.5 [−14.2, −10.8] | 73 | −12.6 |

95% CI by bootstrap over images (2,000 resamples). On the tablet, only the Japanese and US dietitian conditions were run; the BF16 value shown for the tablet rows is that of 4B. Identical: proportion of images whose estimate was the same under both conditions. BF16 server: mean paired difference on the unified reference tables; for 2B, 2–19 responses per condition failed to parse, so the number of pairs is 879–885 on NI887 and 1,112–1,127 on SN1134 (886 and 1,131 for 4B and 8B).

**Table S4. Agreement of the five values between the tablet (iPad Air, MLX Swift) and the desktop computer (MLX), Qwen3-VL-4B at 4 bits.**

| Set | Prompt | Value | n | Identical (%) | Tablet higher | Tablet lower | Mean difference (tablet – desktop computer) | SD of difference |
|---|---|---|---|---|---|---|---|---|
| NI887 | Base | Energy (kcal) | 886 | 83.6 | 78 | 67 | +0.62 | 24.01 |
| NI887 | Base | Protein (g) | 886 | 81.4 | 85 | 80 | +0.02 | 1.93 |
| NI887 | Base | Fat (g) | 886 | 79.7 | 90 | 90 | +0.02 | 2.19 |
| NI887 | Base | Carbohydrate (g) | 886 | 75.4 | 112 | 106 | +0.05 | 3.76 |
| NI887 | Base | Salt equivalent (g) | 886 | 84.2 | 76 | 64 | +0.04 | 0.65 |
| NI887 | Japanese dietitian | Energy (kcal) | 886 | 84.3 | 71 | 68 | +0.05 | 21.30 |
| NI887 | Japanese dietitian | Protein (g) | 886 | 82.1 | 81 | 78 | +0.01 | 1.74 |
| NI887 | Japanese dietitian | Fat (g) | 886 | 78.7 | 89 | 100 | +0.11 | 4.49 |
| NI887 | Japanese dietitian | Carbohydrate (g) | 886 | 74.2 | 117 | 112 | +0.03 | 3.57 |
| NI887 | Japanese dietitian | Salt equivalent (g) | 886 | 82.3 | 87 | 70 | +0.02 | 0.46 |
| NI887 | US dietitian | Energy (kcal) | 886 | 83.4 | 86 | 61 | +1.16 | 21.42 |
| NI887 | US dietitian | Protein (g) | 886 | 78.9 | 96 | 91 | +0.04 | 1.86 |
| NI887 | US dietitian | Fat (g) | 886 | 75.4 | 123 | 95 | +0.14 | 2.09 |
| NI887 | US dietitian | Carbohydrate (g) | 886 | 72.0 | 127 | 121 | +0.19 | 3.48 |
| NI887 | US dietitian | Salt equivalent (g) | 886 | 79.8 | 105 | 74 | +0.03 | 0.44 |
| SN1134 | Base | Energy (kcal) | 1131 | 86.5 | 62 | 91 | −1.55 | 23.26 |
| SN1134 | Base | Protein (g) | 1131 | 86.9 | 73 | 75 | −0.07 | 1.59 |
| SN1134 | Base | Fat (g) | 1131 | 83.4 | 87 | 101 | −0.11 | 2.03 |
| SN1134 | Base | Carbohydrate | 1131 | 77.2 | 131 | 127 | +0.01 | 5.03 |

| Set | Prompt | Value | n | Identical (%) | Tablet higher | Tablet lower | Mean difference (tablet − desktop computer) | SD of difference |
|---|---|---|---|---|---|---|---|---|
| | | (g) | | | | | | |
| SN1134 | Base | Salt equivalent (g) | 1131 | 85.8 | 85 | 76 | +0.02 | 0.65 |
| SN1134 | Japanese dietitian | Energy (kcal) | 1131 | 87.1 | 55 | 91 | −2.18 | 20.39 |
| SN1134 | Japanese dietitian | Protein (g) | 1131 | 89.1 | 64 | 59 | +0.00 | 1.23 |
| SN1134 | Japanese dietitian | Fat (g) | 1131 | 84.7 | 79 | 94 | −0.19 | 3.23 |
| SN1134 | Japanese dietitian | Carbohydrate (g) | 1131 | 80.4 | 92 | 130 | −0.28 | 3.45 |
| SN1134 | Japanese dietitian | Salt equivalent (g) | 1131 | 90.6 | 62 | 44 | +0.02 | 0.39 |
| SN1134 | US dietitian | Energy (kcal) | 1131 | 86.6 | 64 | 87 | −1.56 | 21.45 |
| SN1134 | US dietitian | Protein (g) | 1131 | 88.4 | 62 | 69 | −0.04 | 1.37 |
| SN1134 | US dietitian | Fat (g) | 1131 | 84.7 | 75 | 98 | −0.26 | 4.67 |
| SN1134 | US dietitian | Carbohydrate (g) | 1131 | 75.5 | 123 | 154 | −0.38 | 4.71 |
| SN1134 | US dietitian | Salt equivalent (g) | 1131 | 91.1 | 51 | 50 | +0.00 | 0.30 |

Differences are in the unit of each value. The distribution of the absolute differences is given in Table S5.

### Table S5. Distribution of the absolute differences between the tablet and the desktop computer (Qwen3-VL-4B, 4-bit, base condition), in the unit of each value.

| Set | Value | n | Identical (%) | Discordant n | Median \|diff\| (discordant) | 95th pct \|diff\| (all) | Max \|diff\| | \|diff\| > 20% of reference (%) |
|---|---|---|---|---|---|---|---|---|
| NI887 | Energy (kcal) | 886 | 83.6 | 145 | 50 | 50 | 100 | 11.2 |
| NI887 | Protein (g) | 886 | 81.4 | 165 | 5 | 5 | 13 | 16.7 |
| NI887 | Fat (g) | 886 | 79.7 | 180 | 3 | 5 | 20 | 17.3 |
| NI887 | Carbohydrate (g) | 886 | 75.4 | 218 | 5 | 10 | 30 | 18.7 |
| NI887 | Salt equivalent (g) | 886 | 84.2 | 140 | 1 | 2 | 3 | 15.7 |
| SN1134 | Energy (kcal) | 1,131 | 86.5 | 153 | 50 | 50 | 120 | 6.5 |
| SN1134 | Protein (g) | 1,131 | 86.9 | 148 | 3 | 5 | 10 | 9.1 |
| SN1134 | Fat (g) | 1,131 | 83.4 | 188 | 5 | 5 | 20 | 12.6 |
| SN1134 | Carbohydrate (g) | 1,131 | 77.2 | 258 | 5 | 10 | 55 | 12.1 |
| SN1134 | Salt equivalent (g) | 1,131 | 85.8 | 161 | 1 | 2 | 5 | 14.1 |

Discordant: images whose value differed between the two platforms. For energy, the difference exceeded 50 kcal for 36 NI887 images (4.1%) and 43 SN1134 images (3.8%) and 100 kcal for 0 and 3 images.

### Table S6. Accuracy of the five values by model size (Qwen3-VL 2B, 4B and 8B; desktop computer, 4-bit, base condition).

| Set | Model | Value | n | Bias | MedAE | MedAPE (%) | Within ±20% (%) | ρ | r |
|---|---|---|---|---|---|---|---|---|---|
| NI887 | 2B | Energy (kcal) | 886 | +19.0 | 93.4 | 43.0 | 25 | 0.70 | 0.70 |
| NI887 | 2B | Protein (g) | 884 | −1.2 | 2.9 | 41.9 | 22 | 0.76 | 0.72 |
| NI887 | 2B | Fat (g) | 878 | −0.8 | 3.4 | 53.5 | 18 | 0.72 | 0.68 |
| NI887 | 2B | Carbohydrate (g) | 884 | +2.1 | 11.9 | 69.0 | 14 | 0.54 | 0.56 |
| NI887 | 2B | Salt equivalent (g) | 835 | −0.10 | 0.60 | 45.9 | 23 | 0.41 | 0.18 |

| Set | Model | Value | n | Bias | MedAE | MedAPE (%) | Within ±20% (%) | ρ | r |
|---|---|---|---|---|---|---|---|---|---|
| NI887 | 4B | Energy (kcal) | 886 | +1.1 | 66.4 | 37.0 | 28 | 0.76 | 0.73 |
| NI887 | 4B | Protein (g) | 884 | +2.5 | 3.3 | 46.0 | 24 | 0.79 | 0.76 |
| NI887 | 4B | Fat (g) | 878 | +0.7 | 3.7 | 61.5 | 17 | 0.73 | 0.67 |
| NI887 | 4B | Carbohydrate (g) | 884 | +3.1 | 10.7 | 66.0 | 15 | 0.66 | 0.64 |
| NI887 | 4B | Salt equivalent (g) | 835 | +1.11 | 1.27 | 100.0 | 12 | 0.52 | 0.30 |
| NI887 | 8B | Energy (kcal) | 886 | +39.0 | 85.3 | 46.5 | 24 | 0.78 | 0.70 |
| NI887 | 8B | Protein (g) | 884 | +5.6 | 5.1 | 74.8 | 17 | 0.79 | 0.74 |
| NI887 | 8B | Fat (g) | 878 | +2.6 | 4.6 | 66.4 | 18 | 0.71 | 0.66 |
| NI887 | 8B | Carbohydrate (g) | 884 | +1.9 | 9.8 | 63.8 | 18 | 0.67 | 0.62 |
| NI887 | 8B | Salt equivalent (g) | 835 | +1.25 | 1.38 | 103.3 | 11 | 0.50 | 0.30 |
| SN1134 | 2B | Energy (kcal) | 1131 | −37.3 | 106.3 | 38.1 | 27 | 0.71 | 0.64 |
| SN1134 | 2B | Protein (g) | 1113 | −7.1 | 3.8 | 55.5 | 13 | 0.82 | 0.76 |
| SN1134 | 2B | Fat (g) | 1095 | −4.3 | 4.9 | 57.5 | 15 | 0.69 | 0.58 |
| SN1134 | 2B | Carbohydrate (g) | 1107 | +0.3 | 13.9 | 44.9 | 22 | 0.51 | 0.35 |
| SN1134 | 2B | Salt equivalent (g) | 1119 | −0.22 | 0.74 | 69.4 | 14 | 0.69 | 0.42 |
| SN1134 | 4B | Energy (kcal) | 1131 | −52.0 | 84.2 | 36.1 | 27 | 0.78 | 0.71 |
| SN1134 | 4B | Protein (g) | 1113 | −2.7 | 3.6 | 44.1 | 23 | 0.85 | 0.76 |
| SN1134 | 4B | Fat (g) | 1095 | −2.7 | 4.8 | 61.9 | 16 | 0.73 | 0.60 |
| SN1134 | 4B | Carbohydrate (g) | 1107 | +1.2 | 12.1 | 41.6 | 26 | 0.66 | 0.59 |
| SN1134 | 4B | Salt equivalent (g) | 1119 | +0.29 | 0.51 | 68.4 | 16 | 0.81 | 0.67 |
| SN1134 | 8B | Energy (kcal) | 1131 | +0.6 | 95.2 | 36.2 | 31 | 0.78 | 0.70 |
| SN1134 | 8B | Protein (g) | 1113 | −0.3 | 4.3 | 45.6 | 23 | 0.86 | 0.76 |
| SN1134 | 8B | Fat (g) | 1095 | −0.8 | 5.4 | 55.6 | 18 | 0.73 | 0.60 |
| SN1134 | 8B | Carbohydrate (g) | 1107 | +2.0 | 11.2 | 38.8 | 28 | 0.70 | 0.63 |
| SN1134 | 8B | Salt equivalent (g) | 1119 | +0.62 | 0.76 | 99.7 | 14 | 0.81 | 0.66 |

Bias and median absolute error (MedAE) are in the unit of each value. Images with a reference value of 0 for the value concerned were excluded.

**Table S7. Accuracy of the five values with the food weight supplied (NI887, desktop computer, 4-bit): true weight for 2B, 4B and 8B, and 11 declared-weight conditions for 4B.**

| Model | Declared weight | Value | n | Bias | MedAPE (%) | ΔMedAPE vs base [95% CI] | ρ |
|---|---|---|---|---|---|---|---|
| 2B | None (base) | Energy (kcal) | 886 | +19.0 | 43.0 | — | 0.70 |
| 2B | None (base) | Protein (g) | 884 | −1.2 | 41.9 | — | 0.76 |
| 2B | None (base) | Fat (g) | 878 | −0.8 | 53.5 | — | 0.72 |
| 2B | None (base) | Carbohydrate (g) | 884 | +2.1 | 69.0 | — | 0.54 |
| 2B | None (base) | Salt equivalent (g) | 835 | −0.10 | 45.9 | — | 0.41 |
| 2B | W × 1.0 | Energy (kcal) | 886 | −30.3 | 38.3 | −4.7 [−8.8, −1.7] | 0.79 |
| 2B | W × 1.0 | Protein (g) | 884 | −3.1 | 41.2 | −0.6 [−3.4, +2.7] | 0.82 |

| Model | Declared weight | Value | n | Bias | MedAPE (%) | ΔMedAPE vs base [95% CI] | ρ |
|---|---|---|---|---|---|---|---|
| 2B | W × 1.0 | Fat (g) | 878 | −3.2 | 56.8 | +3.3 [−0.1, +7.9] | 0.73 |
| 2B | W × 1.0 | Carbohydrate (g) | 884 | +0.8 | 59.4 | −9.6 [−16.7, −4.5] | 0.53 |
| 2B | W × 1.0 | Salt equivalent (g) | 835 | −0.33 | 51.7 | +5.8 [+1.6, +10.5] | 0.40 |
| 4B | None (base) | Energy (kcal) | 886 | +1.1 | 37.0 | — | 0.76 |
| 4B | None (base) | Protein (g) | 884 | +2.5 | 46.0 | — | 0.79 |
| 4B | None (base) | Fat (g) | 878 | +0.7 | 61.5 | — | 0.73 |
| 4B | None (base) | Carbohydrate (g) | 884 | +3.1 | 66.0 | — | 0.66 |
| 4B | None (base) | Salt equivalent (g) | 835 | +1.11 | 100.0 | — | 0.52 |
| 4B | W × 1.0 | Energy (kcal) | 886 | +9.8 | 32.5 | −4.5 [−7.3, −2.1] | 0.84 |
| 4B | W × 1.0 | Protein (g) | 884 | +2.7 | 41.7 | −4.2 [−8.1, −0.1] | 0.85 |
| 4B | W × 1.0 | Fat (g) | 878 | +1.1 | 63.3 | +1.7 [−3.3, +5.7] | 0.76 |
| 4B | W × 1.0 | Carbohydrate (g) | 884 | +2.6 | 63.7 | −2.3 [−7.1, +2.8] | 0.74 |
| 4B | W × 1.0 | Salt equivalent (g) | 835 | +0.61 | 66.8 | −33.2 [−43.0, −23.6] | 0.59 |
| 4B | W × 0.5 | Energy (kcal) | 886 | −43.6 | 34.9 | −2.1 [−5.6, +0.5] | 0.83 |
| 4B | W × 0.5 | Protein (g) | 884 | −0.4 | 35.5 | −10.5 [−14.6, −5.6] | 0.85 |
| 4B | W × 0.5 | Fat (g) | 878 | −1.1 | 63.6 | +2.1 [−3.5, +6.6] | 0.77 |
| 4B | W × 0.5 | Carbohydrate (g) | 884 | −3.8 | 49.7 | −16.3 [−21.5, −10.2] | 0.74 |
| 4B | W × 0.5 | Salt equivalent (g) | 835 | +0.08 | 51.1 | −48.9 [−60.6, −38.2] | 0.59 |
| 4B | W × 0.75 | Energy (kcal) | 886 | −11.6 | 30.8 | −6.2 [−9.2, −3.5] | 0.84 |
| 4B | W × 0.75 | Protein (g) | 884 | +1.4 | 37.6 | −8.4 [−12.5, −4.2] | 0.85 |
| 4B | W × 0.75 | Fat (g) | 878 | +0.2 | 62.1 | +0.5 [−4.7, +4.7] | 0.76 |
| 4B | W × 0.75 | Carbohydrate (g) | 884 | +0.1 | 54.6 | −11.4 [−15.8, −5.9] | 0.74 |
| 4B | W × 0.75 | Salt equivalent (g) | 835 | +0.40 | 58.0 | −42.0 [−51.8, −31.4] | 0.58 |
| 4B | W × 0.9 | Energy (kcal) | 886 | +1.9 | 30.7 | −6.2 [−9.0, −3.7] | 0.84 |
| 4B | W × 0.9 | Protein (g) | 884 | +2.2 | 38.6 | −7.4 [−11.3, −3.1] | 0.85 |
| 4B | W × 0.9 | Fat (g) | 878 | +0.8 | 60.7 | −0.8 [−6.0, +3.9] | 0.77 |
| 4B | W × 0.9 | Carbohydrate (g) | 884 | +1.5 | 57.6 | −8.4 [−12.6, −2.6] | 0.74 |
| 4B | W × 0.9 | Salt equivalent (g) | 835 | +0.53 | 62.9 | −37.1 [−46.9, −26.9] | 0.59 |
| 4B | W × 1.1 | Energy (kcal) | 886 | +16.1 | 33.3 | −3.7 [−6.4, −1.3] | 0.84 |
| 4B | W × 1.1 | Protein (g) | 884 | +3.1 | 43.3 | −2.6 [−6.6, +1.7] | 0.85 |
| 4B | W × 1.1 | Fat (g) | 878 | +1.5 | 63.3 | +1.8 [−3.1, +6.1] | 0.77 |
| 4B | W × 1.1 | Carbohydrate (g) | 884 | +3.8 | 67.3 | +1.3 [−3.5, +8.4] | 0.73 |
| 4B | W × 1.1 | Salt equivalent (g) | 835 | +0.67 | 68.9 | −31.1 [−39.3, −21.3] | 0.59 |
| 4B | W × 1.25 | Energy (kcal) | 886 | +25.4 | 34.7 | −2.2 [−5.5, −0.2] | 0.85 |
| 4B | W × 1.25 | Protein (g) | 884 | +3.7 | 46.6 | +0.6 [−3.5, +4.9] | 0.86 |
| 4B | W × 1.25 | Fat (g) | 878 | +1.9 | 63.5 | +1.9 [−2.7, +6.5] | 0.77 |
| 4B | W × 1.25 | Carbohydrate (g) | 884 | +5.5 | 69.9 | +3.9 [−2.2, +14.7] | 0.74 |
| 4B | W × 1.25 | Salt equivalent (g) | 835 | +0.79 | 74.0 | −26.0 [−35.0, −16.6] | 0.59 |
| 4B | W × 1.5 | Energy (kcal) | 886 | +40.6 | 38.0 | +1.0 [−2.0, +4.5] | 0.84 |
| 4B | W × 1.5 | Protein (g) | 884 | +4.7 | 51.6 | +5.6 [+1.8, +10.0] | 0.85 |
| 4B | W × 1.5 | Fat (g) | 878 | +2.5 | 65.5 | +4.0 [−0.4, +8.5] | 0.76 |
| 4B | W × 1.5 | Carbohydrate (g) | 884 | +8.0 | 86.2 | +20.2 [+11.7, +27.8] | 0.74 |
| 4B | W × 1.5 | Salt equivalent (g) | 835 | +0.93 | 85.2 | −14.8 [−23.9, −5.9] | 0.58 |
| 4B | W × 2.0 | Energy (kcal) | 886 | +66.4 | 46.0 | +9.0 [+5.2, | 0.84 |

| Model | Declared weight | Value | n | Bias | MedAPE (%) | ΔMedAPE vs base [95% CI] | ρ |
|---|---|---|---|---|---|---|---|
| | | | | | | +14.4] | |
| 4B | W × 2.0 | Protein (g) | 884 | +6.1 | 64.0 | +18.0 [+13.5, +23.0] | 0.85 |
| 4B | W × 2.0 | Fat (g) | 878 | +3.6 | 71.8 | +10.3 [+4.2, +14.6] | 0.76 |
| 4B | W × 2.0 | Carbohydrate (g) | 884 | +13.0 | 103.6 | +37.6 [+30.8, +51.8] | 0.73 |
| 4B | W × 2.0 | Salt equivalent (g) | 835 | +1.15 | 99.7 | −0.3 [−10.2, +7.7] | 0.58 |
| 4B | W + 150 g | Energy (kcal) | 886 | +79.3 | 56.1 | +19.1 [+14.8, +25.7] | 0.79 |
| 4B | W + 150 g | Protein (g) | 884 | +6.4 | 69.1 | +23.1 [+18.3, +30.5] | 0.81 |
| 4B | W + 150 g | Fat (g) | 878 | +4.0 | 73.1 | +11.5 [+6.1, +19.3] | 0.73 |
| 4B | W + 150 g | Carbohydrate (g) | 884 | +13.2 | 121.7 | +55.7 [+41.2, +69.7] | 0.72 |
| 4B | W + 150 g | Salt equivalent (g) | 835 | +1.23 | 103.9 | +3.9 [−3.2, +15.2] | 0.52 |
| 4B | W + 300 g | Energy (kcal) | 886 | +115.7 | 74.9 | +37.9 [+31.1, +47.9] | 0.76 |
| 4B | W + 300 g | Protein (g) | 884 | +8.8 | 100.0 | +54.0 [+45.3, +62.3] | 0.79 |
| 4B | W + 300 g | Fat (g) | 878 | +5.6 | 79.7 | +18.1 [+10.5, +23.6] | 0.72 |
| 4B | W + 300 g | Carbohydrate (g) | 884 | +22.1 | 167.0 | +101.0 [+86.9, +115.0] | 0.68 |
| 4B | W + 300 g | Salt equivalent (g) | 835 | +1.55 | 130.0 | +30.0 [+17.4, +41.0] | 0.47 |
| 4B | W + 450 g | Energy (kcal) | 886 | +147.4 | 85.8 | +48.9 [+39.1, +60.2] | 0.73 |
| 4B | W + 450 g | Protein (g) | 884 | +10.7 | 116.7 | +70.8 [+62.1, +79.0] | 0.78 |
| 4B | W + 450 g | Fat (g) | 878 | +6.7 | 82.8 | +21.3 [+13.9, +28.5] | 0.72 |
| 4B | W + 450 g | Carbohydrate (g) | 884 | +27.7 | 201.5 | +135.5 [+119.7, +157.9] | 0.67 |
| 4B | W + 450 g | Salt equivalent (g) | 835 | +1.83 | 147.9 | +47.9 [+35.7, +59.9] | 0.47 |
| 4B | W rounded to 50 g | Energy (kcal) | 886 | +11.8 | 31.8 | −5.2 [−8.3, −2.3] | 0.84 |
| 4B | W rounded to 50 g | Protein (g) | 884 | +2.7 | 42.8 | −3.1 [−7.4, +1.1] | 0.85 |
| 4B | W rounded to 50 g | Fat (g) | 878 | +0.8 | 62.1 | +0.6 [−5.4, +4.8] | 0.76 |
| 4B | W rounded to 50 g | Carbohydrate (g) | 884 | +3.1 | 63.5 | −2.5 [−7.2, +3.3] | 0.74 |
| 4B | W rounded to 50 g | Salt equivalent (g) | 835 | +0.81 | 80.5 | −19.5 [−29.6, −12.2] | 0.58 |
| 8B | None (base) | Energy (kcal) | 886 | +39.0 | 46.5 | — | 0.78 |
| 8B | None (base) | Protein (g) | 884 | +5.6 | 74.8 | — | 0.79 |
| 8B | None (base) | Fat (g) | 878 | +2.6 | 66.4 | — | 0.71 |
| 8B | None (base) | Carbohydrate (g) | 884 | +1.9 | 63.8 | — | 0.67 |
| 8B | None (base) | Salt equivalent (g) | 835 | +1.25 | 103.3 | — | 0.50 |
| 8B | W × 1.0 | Energy (kcal) | 886 | +64.0 | 48.2 | +1.8 [−2.7, +6.6] | 0.82 |
| 8B | W × 1.0 | Protein (g) | 884 | +5.1 | 67.5 | −7.3 [−15.3, +0.0] | 0.85 |
| 8B | W × 1.0 | Fat (g) | 878 | +3.3 | 66.4 | −0.1 [−6.7, +5.9] | 0.74 |
| 8B | W × 1.0 | Carbohydrate (g) | 884 | +5.7 | 66.9 | +3.1 [−3.4, +8.2] | 0.70 |
| 8B | W × 1.0 | Salt equivalent (g) | 835 | +1.15 | 94.9 | −8.3 [−22.6, −1.1] | 0.59 |

W, true weight of the food. ΔMedAPE: difference from the base condition of the same model, with 95% CI by paired bootstrap over images (2,000 resamples).
